# A comprehensive framework to estimate the frequency, duration and risk factors for diagnostic delays using simulation-based methods

**DOI:** 10.1101/2021.10.22.21265386

**Authors:** Aaron C Miller, Joseph E Cavanaugh, Alan T Arakkal, Scott H Koeneman, Philip M Polgreen

**Author notes:** Corresponding Author: Aaron C Miller, ^1^Department of Internal Medicine, Roy J. and Lucille A. Carver College of Medicine, University of Iowa, Iowa City, Iowa, 52242.

## Abstract

The incidence of diagnostic delays is unknown for many diseases and particular healthcare settings. Many existing methods to identify diagnostic delays are resource intensive or inapplicable to various diseases or settings. In this paper we propose a comprehensive framework to estimate the frequency of missed diagnostic opportunities for a given disease using real-world longitudinal data sources. We start by providing a conceptual model of the disease-diagnostic, data-generating process. We then propose a simulation-based method to estimate measures of the frequency of missed diagnostic opportunities and duration of delays. This approach is specifically designed to identify missed diagnostic opportunities based on signs and symptoms that occur prior to an initial diagnosis, while accounting for expected patterns of healthcare that may appear as coincidental symptoms. Three different simulation algorithms are described for implementing this approach. We summarize estimation procedures that may be used to parameterize the simulation. Finally, we apply our approach to the diseases of tuberculosis, acute myocardial infarction, and stroke and evaluate the estimated frequency and duration of diagnostic delays for these diseases. Our approach can be customized to fit a range of disease and we summarize how the choice of simulation algorithm may impact the resulting estimates.

## Background

Diagnostic errors are a major contributor to morbidity, mortality and excess healthcare costs.^1,2^ One of the most common types of diagnostic errors are diagnostic delays. For many diseases, timely diagnosis is essential for effective treatment, and for some diseases even minimal delays may significantly increase risk of patient harm.^3,4^ Identifying cases where diagnostic delays have occurred is a critical first step in studying the causes and consequences of diagnostic delays and for developing interventions to prevent delays. However, for many diseases and settings, the incidence of diagnostic delays is unknown or challenging to estimate.^5,6^

Historically, a number of approaches have been used to study diagnostic delays; these include retrospective reviews of medical records, autopsy studies, analysis of malpractice claims, and patient or clinician surveys. ^5,7^ These approaches are highly informative, but have a number of limitations. For example, chart reviews are labor intensive, and have been primarily focused on single hospitals or health systems, thus limiting their generalizability. Other approaches, such as studies of autopsy results or malpractice claims may only apply to the most serious cases or diseases. Moreover, many approaches to study diagnostic delays have exclusively focused on hospital records or emergency department settings.^8-13^ Yet, many opportunities to diagnose a disease occur in outpatient clinics,^14,15^ and patient care often occurs across a wide spectrum of disconnected facilities. Thus, longitudinal information spanning a wide variety of healthcare settings and covering a broad patient population is required to fully capture the diverse spectrum of diagnostic delays.

Another limitation of most investigations of diagnostic delays, is that the criteria used to define a diagnostic delay must be specified *a priori*. Typically, expert evaluation must be used to determine the criteria to define a diagnostic delay based on what is known about the natural history of the disease prior to diagnosis. These criteria include validating the index diagnosis, describing the clinical signs and symptoms that indicate the disease was present prior to diagnosis, identifying the types of clinic records (e.g., notes, lab results, diagnostic codes, etc.) necessary to capture signs and symptoms of the disease, and selecting the biologically plausible period of time prior to the index diagnosis where an earlier diagnosis could have occurred. ^*8,9*^ However, if a significant number of diagnostic delays occur among patients with atypical presentation or outside the time period considered, such cases may continue to be missed.

Another significant limitation with specifying criteria for a diagnostic delay in an *a priori* fashion is that some patients may meet criteria defining a delay simply by coincidence, especially if the criteria include common clinical signs or symptoms. For example, patients with tuberculosis may have a history of a cough prior to developing tuberculosis or patients may suffer from back pain prior to developing a spinal abscess. In such cases, symptoms may appear to be attributable to the disease, but are actually unrelated. Including such shared but unrelated common clinical signs and symptoms will lead to overestimation of diagnostic delays. Numerous investigations have relied on algorithms to identify diagnostic delays based on commonly occurring symptom criteria such as cough, fever, pain, headaches, malaise and fatigue^8,9,12,16^, yet only a few have attempted to account for a coincidental or expected occurrence of such symptoms.^17-19^

A growing number of investigators have begun to use longitudinal administrative and EMR-based data to identify diagnostic delays.^11,20-22^ These data allow both inpatient, outpatient or emergency department (ED) records to be used in a “look back” approach, where evidence of a disease (e.g., symptom codes) is identified in visits prior to the definitive diagnosis. For example, visits associated with dizziness may be identified prior to a stroke diagnosis,^9,13^ or cough and fever may be identified prior to a tuberculosis diagnosis.^16,18^ Such visits are then considered potential missed opportunities if they occur during a specified *diagnostic opportunity window* - the time before the initial diagnosis where clinical disease may be present and where a diagnostic delay may occur (e.g., 10-days prior to a stroke diagnosis or 90-days prior to a tuberculosis diagnosis). This “look back” approach has been used to study a variety of diseases, including acute myocardial infarctions, strokes, subarachnoid hemorrhages, abdominal aortic aneurysms and tuberculosis,^8-13,16,17^ and has recently been formalized more broadly as the *SPADE* (Symptom-Disease Pair Analysis of Diagnostic Error) framework.^20^

However, three methodological limitations exist with many of the current approaches to study diagnostic delays using observational data. First, as noted above, some signs and symptoms of disease observed prior to diagnosis will not represent actual diagnostic delays, but rather coincidental events that occur prior to the index diagnosis. Second, applications typically require investigators to pre-specify the period of time (i.e., diagnostic opportunity window) prior to diagnosis when delays would be expected to occur. A window that is too long will tend to overestimate the number of diagnostic delays, while a window too short will lead to underestimates. Third, diagnostic codes for symptoms associated with a diagnosis may be underutilized.^23^ For example, patients with a cough may not receive a diagnostic code for cough and instead be assigned a code for pneumonia or respiratory infection. Relying solely on symptom-based codes would miss these visits, especially with conditions that may first be misdiagnosed as an alternative disease (e.g., pneumonia, asthma, COPD or lung cancer instead of tuberculosis).

The purpose of this paper is to expand upon the existing literature using longitudinal observational data sources to study diagnostic delays, while providing a broad methodological framework to address the limitations highlighted above. Specifically, we describe an approach for estimating the frequency of diagnostic delays at a population-level that starts by detecting the point in time where symptomatic visits, associated with the disease of interest, significantly increase prior to the eventual index diagnosis. We then implement a simulation-based approach to estimate the number of “likely” missed diagnostic opportunities that individual patients experience and the duration of diagnostic delays, which would typically require identification of individual patient delays. Finally, we provide a number of different simulation algorithms, considerations for estimation approaches, and a statistical software package, that allow these methods to be customized to a wide range of diseases. This work expands upon the basic conceptual framework described as SPADE by Liberman et al.^20^ It also builds upon the methodological approach utilized by Waxman et al.^17^ to separate observed and expected trends in symptomatic visits prior to diagnosis. Moreover, this study generalizes the methods that the study authors have previously developed to investigate diagnostic delays associated with tuberculosis ^18^ and herpes simplex encephalitis.^19^

The following sections summarize our approach along with three empirical applications and is organized as follows. We start by describing the conceptual framework behind our simulation approach. Next, we outline the basic simulation framework along with three algorithms that may be considered to implement this approach. We then describe some of the estimation procedures that may be used to obtain the parameters necessary to implement the simulation. We also describe sensitivity analyses that may be considered. We then apply our simulation approach to three diseases where diagnostic delays have been previously investigated using large administrative data sources – tuberculosis, acute myocardial infarction (AMI) and stroke. Finally, we describe how results for this disease may differ across the different simulation approaches and estimation procedures. We conclude by discussing considerations for future investigations.

### Theoretical and Conceptual Framework

We define a missed diagnostic opportunity as a healthcare encounter where signs or symptoms of a disease are present, but where the diagnosis is not made or an incorrect diagnosis is applied. Our methodological framework is based on the following fundamental assumption: *for a given disease, a portion of patients will experience missed diagnostic opportunities prior to the index diagnosis of the disease, and such missed opportunities will be reflected by a greater than expected number of healthcare visits where signs and symptoms of the disease are present*.

To identify potential missed opportunities, we start by computing the number of visits prior to the index diagnosis where signs and symptoms of the disease of interest are present. To do so, we expand upon the *symptom-disease pair* concept in the SPADE framework^20^ to include what we term as “*symptomatically similar diagnosis-disease pairs*” where a symptomatically similar diagnosis (SSD) encompasses not only signs and symptoms or related diagnoses, but also tests or procedures that may suggest the presence of the disease.^20^ SSD-related visits may be identified using diagnosis codes (e.g., ICD-9-CM/ICD-10-CM), procedure codes (CPT or ICD), medication claims or other structured data elements. We generally categorize SSDs into one of three types of events (and this list may be expanded upon based on expert-feedback or biological plausibility):

1. **General symptoms** of the disease of interest. For example, for tuberculosis these may include symptoms such as cough, fever, weight loss, or hemoptysis.
2. **Symptomatically-similar diseases or syndromes** that share similar symptoms to the disease of interest and subsequently may be confused for the disease of interest. For example, with tuberculosis these may include pneumonia, influenza or bronchitis.
3. **Testing, imaging, physical-exam-based diagnoses, or treatments** that are associated with symptoms of the disease of interest. For tuberculosis, these may include factors such as infection testing, chest x-rays, diagnoses of anemia or swollen lymph nodes.

We analyze diagnostic opportunities by evaluating the trend in SSD-related visits prior to the index diagnosis. As an example, Figure 1 depicts SSD-related visits prior to the index tuberculosis diagnosis for 3,371 patients (additional details about this study population and SSD conditions are described below). The x-axis depicts the number of days prior to the index tuberculosis diagnosis, and the y-axis depicts the number of visits that occurred on a given day that had an SSD-related diagnosis. From Figure 1, we see that there is a large visible spike in the number of visits for SSD conditions that might be related to tuberculosis in the time just prior to the initial diagnosis. In this case, the dramatic increase appears to occur around 90-100 days prior to the index diagnosis, a time period consistent with prior investigations for when diagnostic delays for tuberculosis might occur.^16,24^

**Figure 1.**
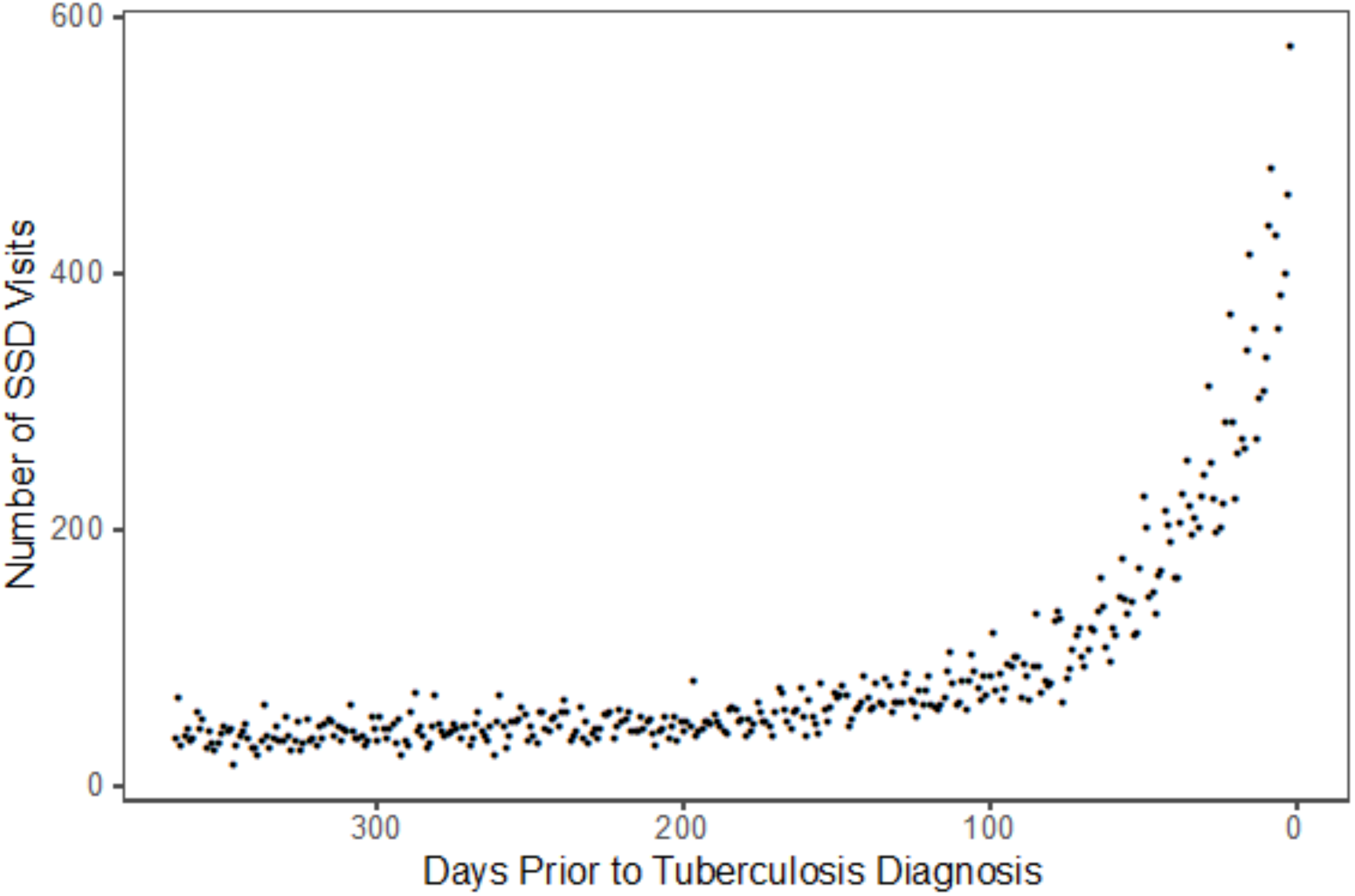
Count of SSD-related visits prior to tuberculosis diagnosis aggregated across all patients with index tuberculosis diagnosis.

The trends presented in Figure 1 have been broadly identified for a wide range of diseases and numerous studies have used this spike in healthcare utilization prior to diagnosis as a marker for diagnostic opportunities.^8,9,17^ This trend depicts two periods of activity: (1) a window just prior to diagnosis where SSD-related visits appear to dramatically increase, which we refer to as the ***diagnostic opportunity window*** and (2) a period further before diagnosis where SSD visits appear to exhibit either a stable or slightly increasing trend. These two periods are highlighted in Figure 2, separated by the dashed-grey line and represent points in time where diagnostic delays are likely to occur (to the right) or unlikely to occur (to the left). The period prior to the diagnostic opportunity window is depicted by a relatively gradual, and near linear, trend in SSD visits. This period may capture risk factors for this disease or the natural history of the disease, but generally does not reflect missed diagnostic opportunities. The increase in visits over time may reflect visits attributable to risk factors for the condition of interest, deteriorating health or surveillance/observation effects (e.g., patient care may cluster in time based on insurance enrollment, patient scheduling convenience or when subsequent visits are for follow-up care or in response to screening).

**Figure 2.**
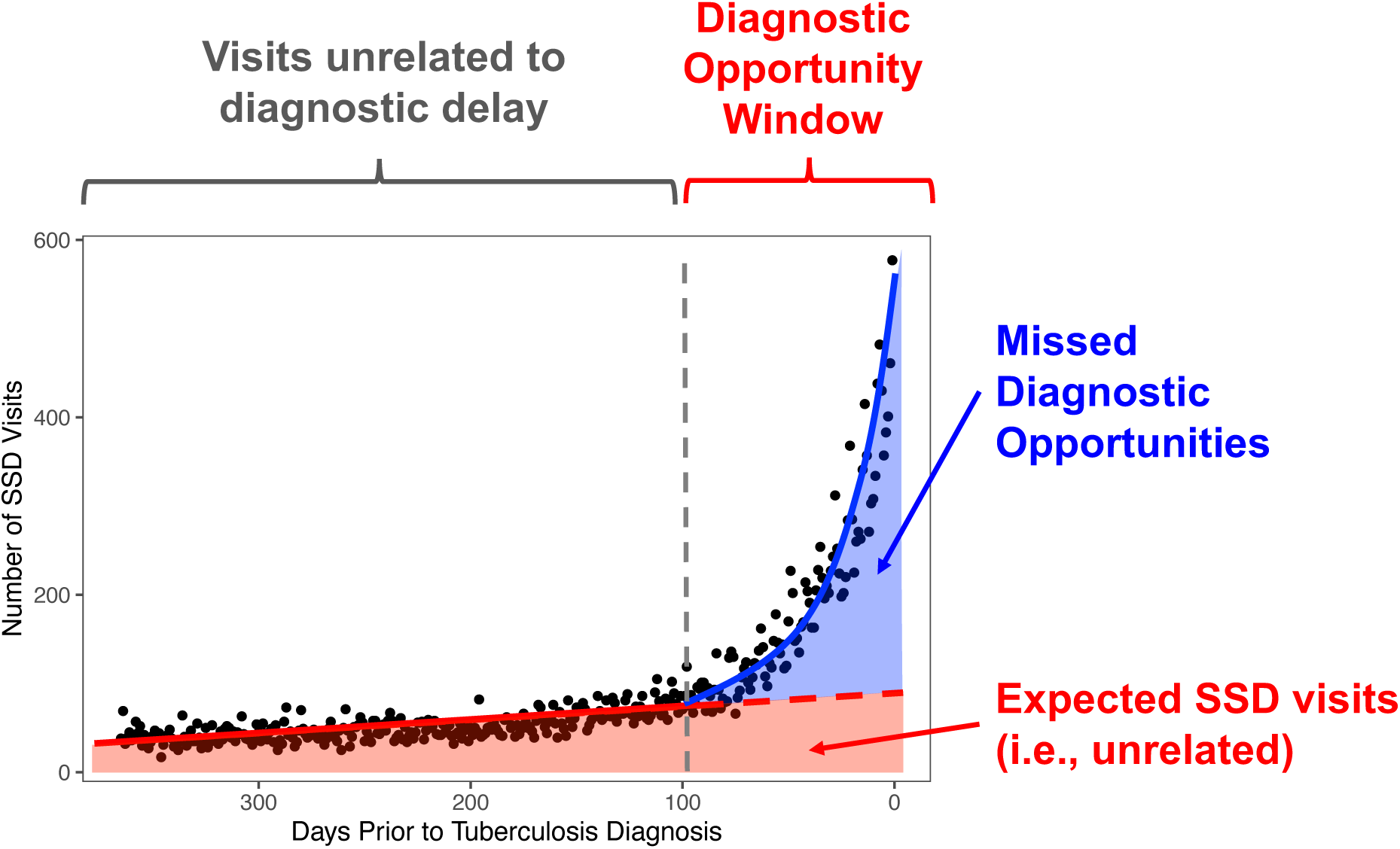
Diagram of conceptual framework representing the number of missed diagnostic opportunities. The diagnostic opportunity window represents the period of time where diagnostic opportunities may occur. The red line depicts the trend in the number of SSD visits that would be expected to occur in absence of diagnostic delays. The blue curve represents the observed trend in SSD visits during the diagnostic opportunity window. The shaded blue region corresponds to the number of missed diagnostic opportunities.

We build upon the distinction between observed and expected trends outlined by Waxman et al. (2018) and our prior empirical work^18,19^ to distinguish missed opportunities from coincidental care. Specifically, the red line in Figure 2, depicts the ***expected*** number of SSD visits – these represent SSD-related visits that would be expected to occur in the absence of diagnostic delays. The solid-red line, reflects the observed trend in SSD-visits prior to the *diagnostic opportunity window*, while the dashed-red line reflects this trend extrapolated to the diagnostic opportunity window. The expected number of visits represents the number of SSD-visits one would expect if the disease of interest were not present. Notice that this extrapolation reflects a type of *case-crossover design*, where the period prior to diagnostic opportunity window is used as a control period to estimate the expected number of visits (if the disease were absent). The visits approximated by the red shaded area, below the *expected* trend line represent the number of SSD visits that would be expected to occur in the absence of diagnostic opportunities.

The blue curve in Figure 2 represents the ***observed*** trend in SSD-visits during the diagnostic opportunity window. The shaded blue area, between the observed and expected trends inside the diagnostic opportunity window, represents the ***excess*** number of SSD-visits. This area roughly approximates the number of visits representing missed diagnostic opportunities. However, some of the visits during the delay opportunity window would also be expected to occur based on trends prior to this period (i.e., visits shaded in red to the right of the dashed-grey line), which are not considered missed opportunities. For example, we would expect some patients with tuberculosis to have had pneumonia within 90 days prior to their tuberculosis diagnosis simply by coincidence.

Given estimates for the observed and expected trends during the diagnostic opportunity window, one can approximate the ***number of missed opportunities*** on a given day by subtracting the number of expected SSD visits from the number of observed SSD visits. (Note: either the true number of observed visits or the estimated trend in observed visits, as depicted by the blue line, may be used in this context.) The observed and expected trends depicted in Figure 2 can be estimated in a variety of ways (e.g., linear or non-linear curves or non-parametric approaches). Once the trends have been estimated the number of missed opportunities on a given day can be computed by subtracting the number of expected SSD visits from the number of observed SSD visits. This and similar approaches using observed and expected visits have been used in prior investigations of diagnostic delays.^17-19^

### Simulation Approach to Identify Likely Missed Opportunities

The above framework may be used to estimate the number of missed opportunities each day during the diagnostic opportunity window. However, because some visits represent *expected* SSD-related visits during the diagnostic opportunity window (i.e., the red shaded region in Figure 2), it is often not possible to directly identify which individual patient visits exactly represent a missed opportunity from observational data alone. Thus, from the estimate of the number of missed opportunities alone we cannot determine: (1) the number of patients who experienced a missed opportunity, (2) the typical duration of diagnostic delays nor (3) the number of missed opportunities that a typical patient experienced. Moreover, it may be challenging to estimate risk factors for experiencing a missed opportunity if individual visits representing a diagnostic opportunity cannot be identified. We refer to these types of measures as *patient-level metrics* associated with diagnostic delays. Our simulation framework is designed to estimate each of these patient-level metrics using a bootstrapping-based approach. We do so by simulating (i.e., randomly selecting) which visits represent a missed opportunity and then computing the individual-patient-level metrics of interest.

Let *m*_*t*_ denote the number of estimated missed opportunities at each day *t* ∈ {*w, w* + 1, …, −2, −1} during the diagnostic opportunity window, where *w* < 0 denotes the point representing the start of the diagnostic opportunity window (see dashed grey line in Figure 2).

Below we describe three different algorithms that may be used to simulate missed visits. In general, these simulation approaches can be described by the following steps. Given estimates for *m*_*t*_ and *cp*, described above, do the following:

1. For each time period in the delay opportunity window, *t* ∈ {*w, w* + 1, …, −2, −1}, randomly draw the estimated number of missed visits *m*_*t*_ and label these as *missed opportunities*.
2. Aggregate all visits and corresponding patients who were drawn to represent a missed opportunity. Compute the number of patients missed, duration of delay (the time between first missed visit and the index date) for each patient, and the number of missed opportunities drawn for each patient.
3. Repeat steps 1 and 2 multiple times.
4. Aggregate results.

The following algorithms expand upon the selection procedure described in step 1 by preferentially drawing patient visits in relation to their perceived probability of representing a delay.

#### Algorithm 1: Independent draws

The first approach draws visits representing *missed opportunities* independent of one another at each time period in the delay opportunity window. A formal description of this algorithm is presented in Figure 3. This represents the simplest way to simulate missed opportunities, but provides no correlation structure between the patients or visits that are selected at subsequent time points. In other words, a patient who is drawn to have a missed opportunity because of symptoms occurring at 21 days before diagnosis would be no more likely to be drawn if they presented with symptoms 14 days prior to diagnosis.

**Figure 3.**
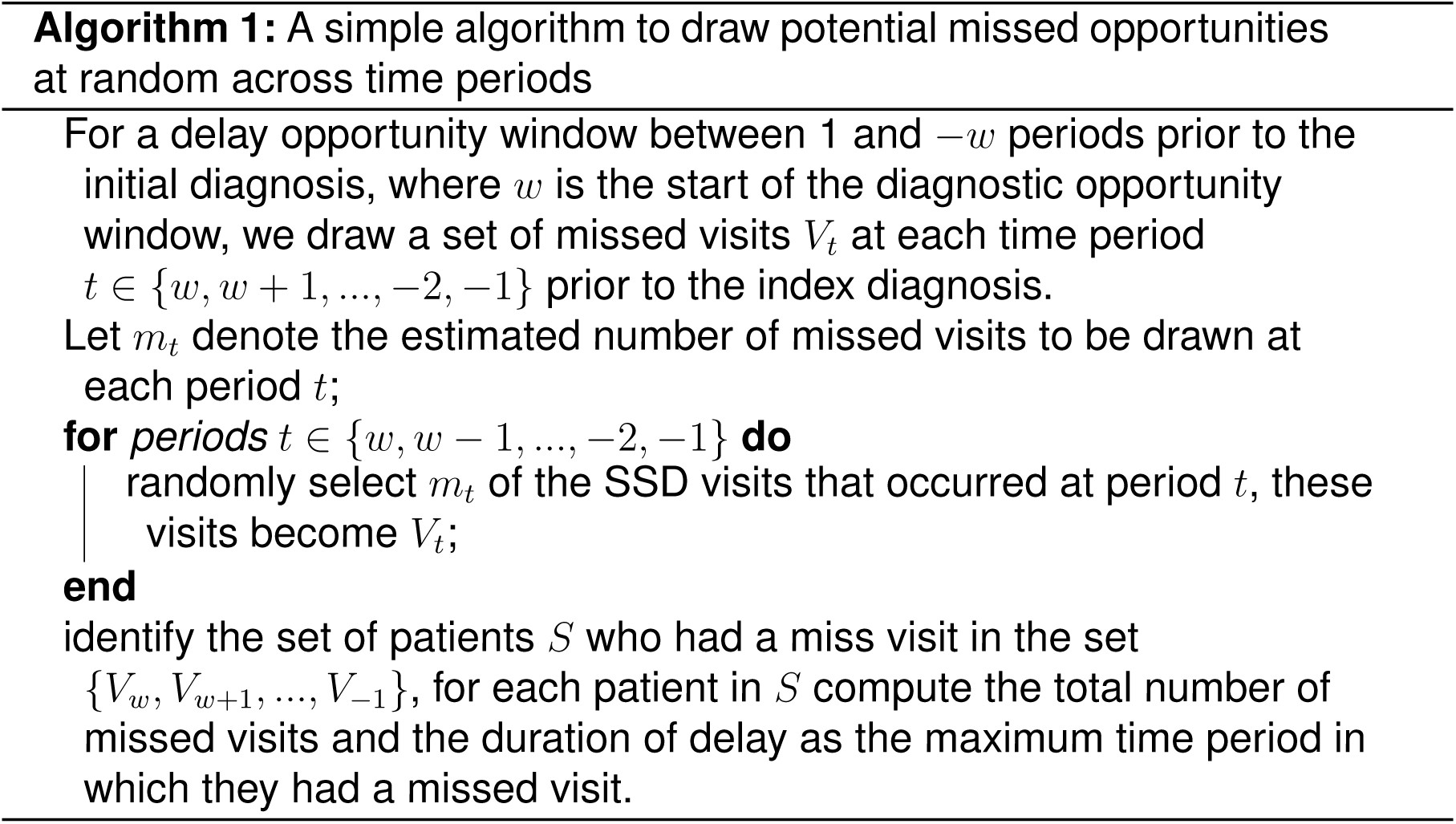
Simple algorithm to simulated missed opportunities using uncorrelated draws.

#### Algorithm 2: Preferential selection of previously drawn cases

In general, SSD visits occurring near the index diagnosis may be more likely to represent a missed opportunity if that patient also experienced a missed opportunity at earlier points before the index diagnosis. For example, if a patient has an SSD visit 15 days prior to the index diagnosis and they also have another SSD visit at 5 days prior to the index, these visits are more likely to be missed opportunities than a single SSD visit. Thus, there may be a desire to draw missed visits in a manner correlated with prior time steps.

Thus, a second approach is to preferentially draw visits from patients who have previously been drawn at earlier time points in the diagnostic opportunity window. Figure 4 presents a formal description of an algorithm to select patient visits in a correlated fashion, where visits are more likely to be selected if the patient also had a missed opportunity at a prior time point. This algorithm introduces a scaling parameter allowing one to define the preference given to selecting patients who have previously been drawn. Specifically, given a scaling parameter *α ϵ* [0,1], *m*_*t*_ * (1 − α) of the visits at time point *t* will be selected from patients with previous missed visits (if available) and *m*_*t*_ * *α* of the visits at time point *t* will be selected from patients not previously drawn to have a missed opportunity. Note that a value of *α* = 0 denotes strict preference to previously drawn patients while *α* = 0.5 denotes equal preference, and *α* = 1 denotes strict preference to patients not drawn at prior time steps. Thus, using this selection procedure and given a set of estimates {*m*_*t*_|*t* ∈ [*cp*, −1]} for missed visits, *α* = 0 will minimize the number of patients with at least one missed opportunity in the diagnostic opportunity window, while *α* = 1 will maximize the number of patients with at least one missed opportunity.

**Figure 4.**
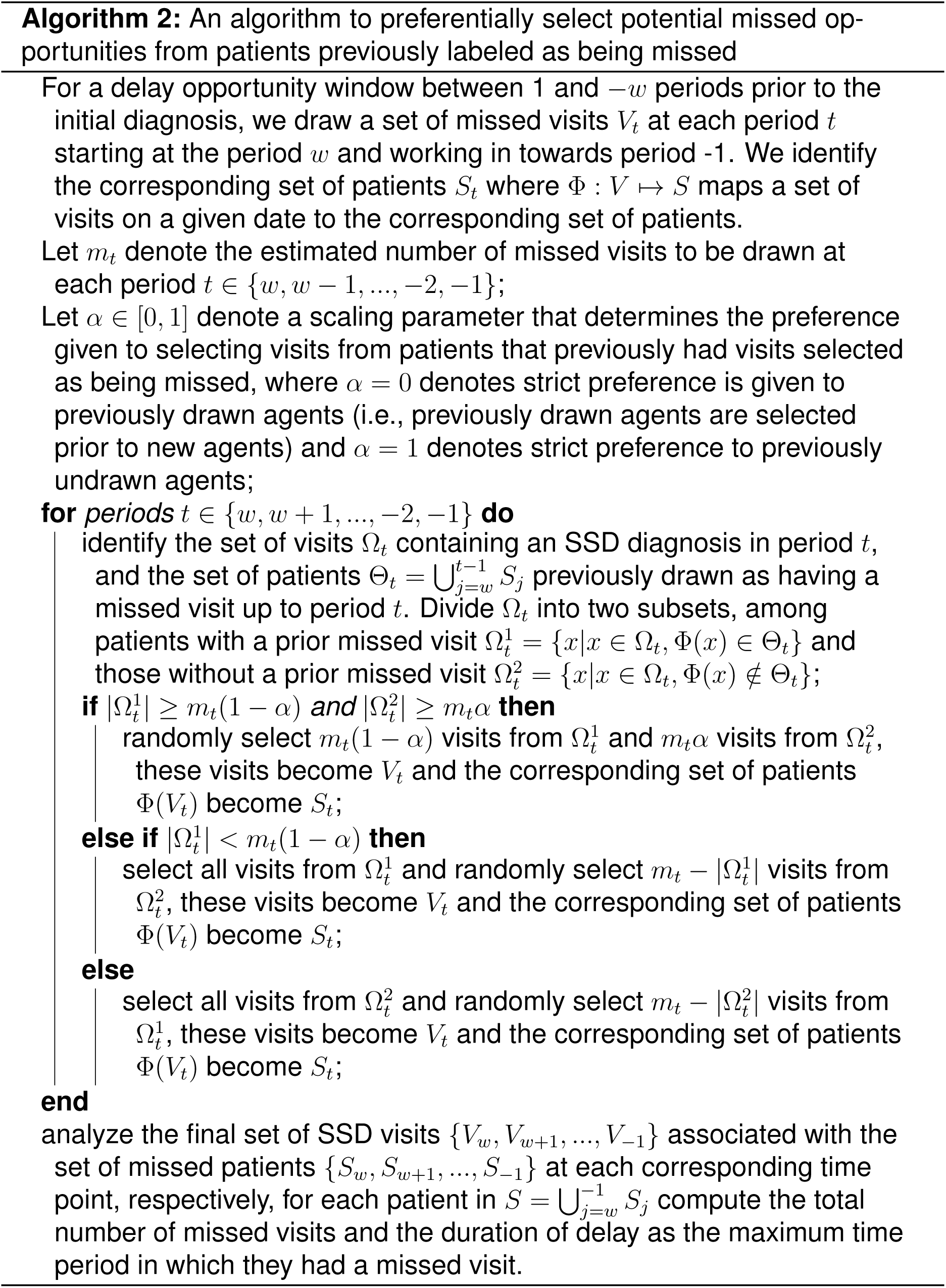
An algorithm to draw patients with preference given to patients previously drawn.

#### Algorithm 3: Generalized algorithm

We may also want to preferentially select visits from patients who are more likely to represent a missed opportunity based on multiple criteria. For example, a patient who has multiple healthcare encounters with unresolved symptoms may be more likely to represent a missed diagnostic opportunity. Similarly, a patient who presents with multiple different symptoms may be more likely to represent a diagnostic delay. Figure 5 presents an example of a more generalized algorithm that allows multiple criteria to be incorporated into the preferential selection criteria. This algorithm incorporates a functional weighting parameter based on the number of times a patient had SSD visits labeled as a missed opportunity and the number of distinct symptoms/SSDs the patient experienced during the current visit or prior visits in the SSD window.

**Figure 5.**
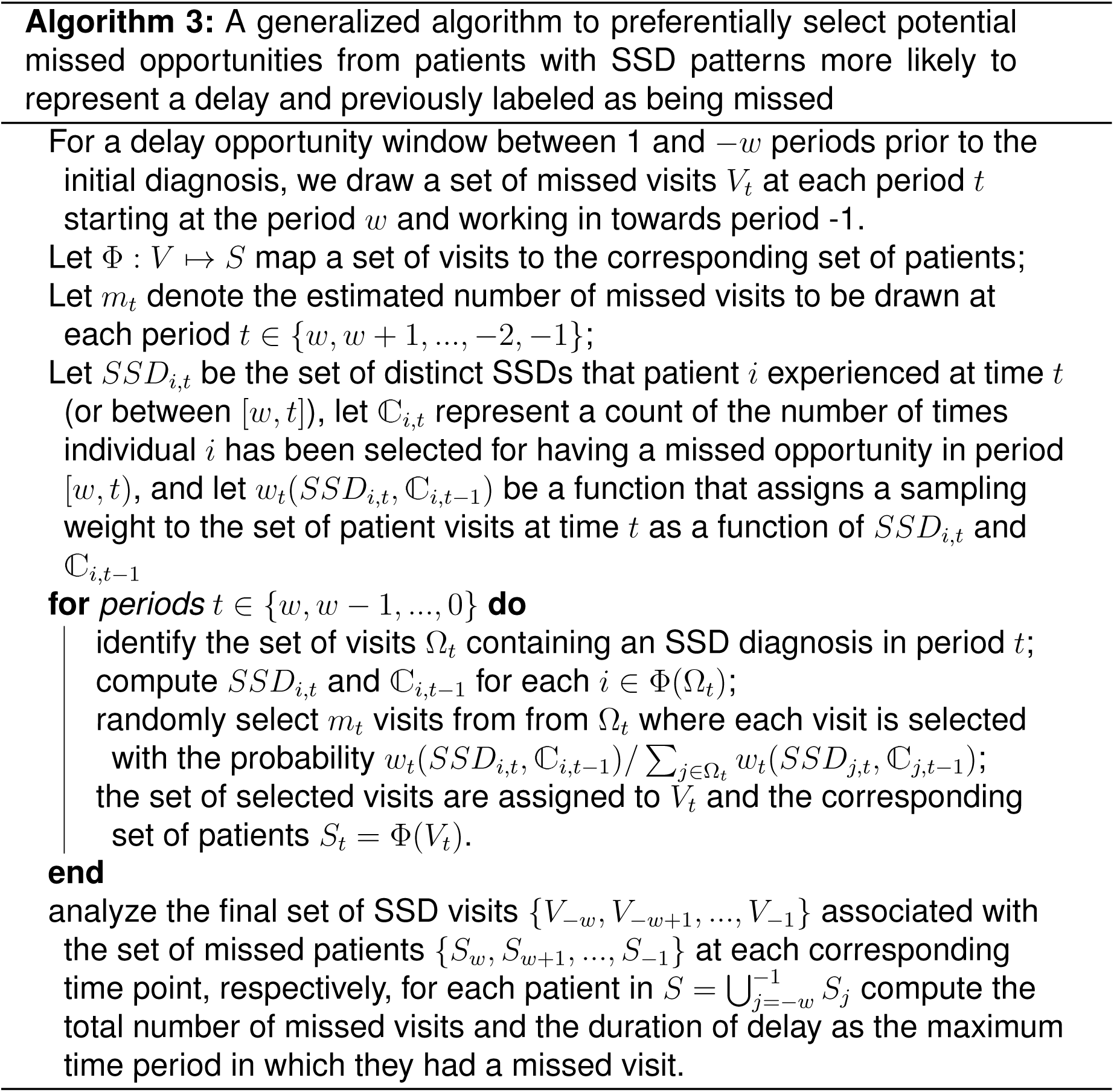
a generalized algorithm to draw patients with preference to previously drawn patients and those with multiple symptoms.

#### R Package

We have developed an R package to implement the above algorithms. This package can be found at https://github.com/aarmiller/delaySim along with installation instructions and tutorial examples.

### Estimating Simulation Parameters

In this section, we discuss approaches to estimate the primary parameters necessary to implement the simulations described above. We begin by describing approaches to estimate the number of missed opportunities each day during the diagnostic opportunity window. We then describe approaches to estimate the bounds of the diagnostic opportunity window. Note: depending on the approach one chooses, these two parameters may be estimated simultaneously.

#### Estimating the expected trend and number of missed opportunities (m_t_)

Let *t* ∈ −*T*, −*T* + 1, …, −2, −1represent time points prior to the index diagnosis, where −*T* represents the maximum amount of time prior to diagnosis that we wish to analyze.

Our goal is then to estimate *y*_*t*_ = *f*(*t*) + *ϵ*_*t*_, where *y*_*t*_ is the number of SSD visits at time *t* over the interval [−*T, w* − 1]. Then using this estimate we extrapolate *ŷ*_*t*_ to the interval [*w*, −1].

Alternatively, we can specify the estimation problem over the entire interval as a piecewise function as follows:

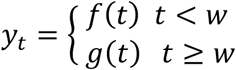

where *f*(*t*) is the trend in SSD visits prior to the diagnostic opportunity window, and *g*(*t*) is the trend in SSD visits during the diagnostic opportunity window. A variety of model fitting approaches may be used to estimate *f*(*t*) and/or *g*(*t*). For example, Figure 6 depicts a case where *f*(*t*) is either a linear function of time (left) or an exponential function (right). Similarly, various time-series modeling approaches may be used to capture temporal aspects of the estimation problem (e.g., periodicity, autocorrelation). In general, we have found that the trend prior to the diagnostic opportunity window (*f*(*t*)) can be roughly approximated by a linear model, while the trend during the diagnostic opportunity window (*g*(*t*)) is typically non-linear.

**Figure 6.**
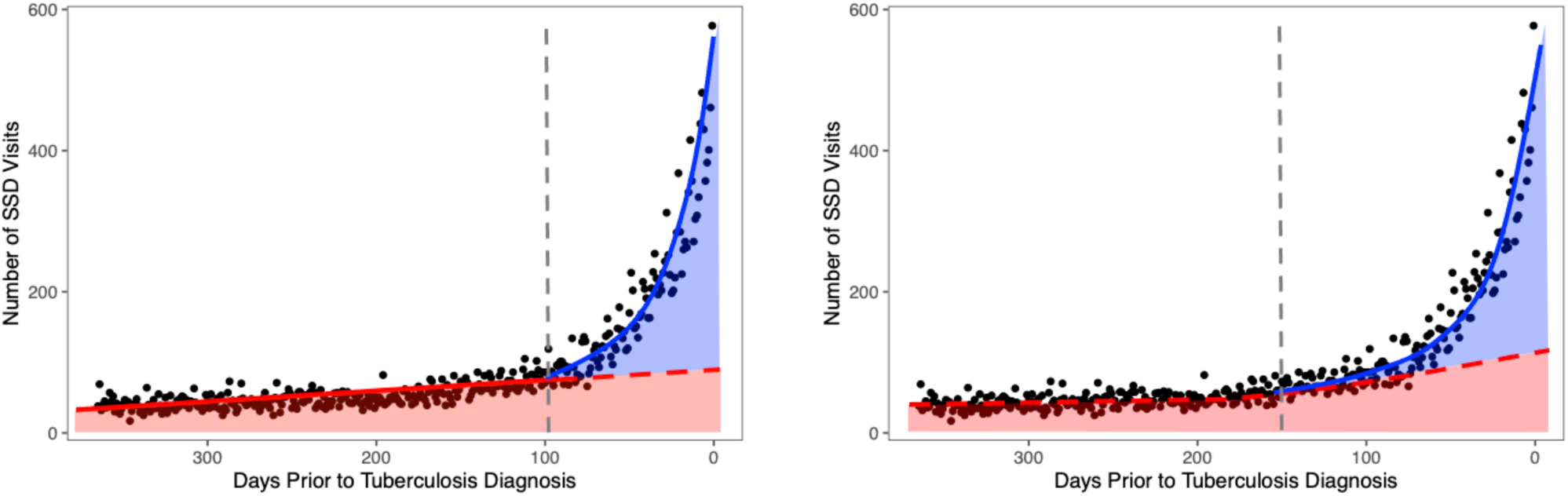
Estimating expected number of visits using linear (left) or exponential (right) curves to represent the expected number of SSD visits.

Once values for 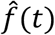 have been obtained, we can compute the number of missed opportunities at a given time point *t* in one of two ways. First, if an explicit value for *ĝ*(*t*) has not been obtained, we can use the observed count at time *t*, such that 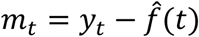. Second, we can use the fitted value for *ĝ*(*t*)such that 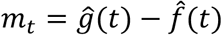. Confidence bounds around the number of missed opportunities may also be computed using appropriate prediction intervals around 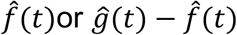.

#### Estimating the bounds of the diagnostic opportunity window (change-point detection)

The lower bound of the diagnostic opportunity window, *w*, represents the cross-over point prior to diagnosis used to delineate the diagnostic opportunity window from the period where the expected pattern of care is estimated. Thus, *w* must be defined prior to calculating *m*_*t*_ as noted above. While this bound on the diagnostic opportunity window may be specified *a priori* based on clinical knowledge, it may also be desirable to estimate this “change point” as part of the analytical process. For example, the maximum duration of diagnostic delays is often the subject of investigation.

One approach for finding this change point is to employ standard change-point finding algorithms to find the optimal value for *cp* = *w* using the trends outlined above. For example, given a parametric specification for *f*(*t*) and *g*(*t*), one approach to find the optimal value *cp* may be achieved by iterating over different values for *cp* and comparing the fitted performance of 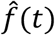 and *ĝ*(*t*) (e.g., by minimizing the Akaike or Bayesian information criterion [AIC/BIC] or maximizing mean squared error [MSE], etc.) Change-point-detection approaches may also be used that do not require explicit specification of functional forms for *f*(*t*) or *g*(*t*) such as the CUSUM method.^25^

Alternatively, if one wishes to use the formula 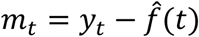, described above, to estimate the number of missed opportunities without estimating *g*(*t*), it may be possible to find *cp* by exploiting the fundamental assumption that *y*_*t*_ > *f*(*t*), ∀ *t* > *cp*. Specifically, we can define the change-point as the point *cp* such that *y*_*t*_ > *ŷ*_*t*_ ∀ *t* > *cp* or using the prediction bound we can define *cp* such that 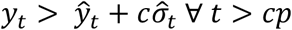, where *c* is a critical value based on the coverage probability. We can then attempt to find a value of *cp*, by choosing an initial guess 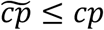 that we believe is outside of the true diagnostic opportunity window, while using the interval 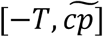 to estimate *f*(*t*). Using this initial guess 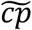, we can then identify the value *cp* in the problem described above. This change-point finding approach is universally applicable regardless of the parameterization of *f*(*t*). Of course the performance of this approach is highly dependent on the initial value 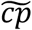, and it is desirable to choose a value as close to the true change point as possible. Note: this approach might be desirable if an upper bound on the feasible diagnostic opportunity window can be clinically justified but where the exact window remains unknown (e.g., diagnostic delays for HSV-encephalitis should never exceed 3 months).

### Applications

We apply our simulation approach to three diseases where a type of “look-back” approach, has been previously applied to study diagnostic delays: stroke, AMI, and tuberculosis. For each of these conditions, we use criteria from these prior studies to identify case patients and the index diagnosis using a large administrative claims dataset. We then compute the number of SSD-related visits each day prior to the diagnosis and use these counts to estimate the number of missed opportunities. Supplementary Table 1 describes the diagnosis codes and sources used to identify each index condition. Supplementary Table 2 describes SSDs used for each condition and their corresponding diagnosis codes. Each of these criteria were selected based on prior investigations of diagnostic delays for these diseases. ^8,9,13,16-18,26-29^

#### Study Population

We used administrative claims data from the IBM Marketscan Commercial Claims Databases from 2001-2017. This database contains longitudinal insurance claims for individuals with employer-sponsored health insurance along with spouses, partners and dependents of the primary enrollee. Over this study period, records are available for over 185 million distinct enrollees. Claims for inpatient, outpatient, emergency department (ED) and prescription medications are provided. For each of the study conditions, we identify the first time an enrollee was diagnosed with the disease of interest and labeled this as the index diagnosis. We exclude children <18 years of age and enrollees that had less than 365 days of continuous enrollment prior to the index diagnosis. Based on prior investigations and clinical plausibility, delays associated with tuberculosis can be expected to exceed a few months while delays for AMI and stroke are typically a month or less. Thus, we used a value of *T* = 365 days for tuberculosis and *T* = 180 days for stroke and AMI.

#### Estimation procedures for simulation parameters

For each condition, we compare three approaches to find the potential change-point marking the start of the diagnostic opportunity window. First, we fit a piecewise linear-cubic model with a linear trend over the interval [−*T, cp* − 1] and a cubic trend over the period [*cp*, −1]. We then iterate over values for *cp* and choose the optimal value based on AIC. Second, we used the CUSUM method to detect the change point over the interval prior to the diagnostic opportunity window beginning at −*T*. For this approach we use a linear model to estimate the expected trend during visits prior to the identified change-point *cp*. Third, we use the prediction bound approach, described above, to identify the point where the observed values are systematically greater than the 95% prediction bound for the expected projections during the diagnostic opportunity window. For this approach, we also use a linear model to estimate the trend in expected SSD visits prior to the change point.

After identifying the change point for each approach, we select the optimal change-point method based on two primary criteria. First, the change point approach should maximize the model fit during the period prior to the diagnostic opportunity window, where we intend to estimate the expected trend in SSD visits that will be extrapolated forward into the diagnostic opportunity window. We characterize this trend by choosing the model that minimizes the mean squared error over the period prior to the diagnostic opportunity window. Second, we evaluate the model performance just prior to the change-point to ensure that the model does not begin overestimating the trend prior to the delay opportunity window. Because the observed trend after the change-point is expected to monotonically increase, if the change-point is set inside the theoretical diagnostic opportunity window we would expect it to result in a negative error as the slope of the expected SSD curve will begin to shift upward. To evaluate this criterion, we evaluate the mean error within 7, 17, and 21 days prior to the change-point, and choose the model with a mean error consistently nearest to zero.

After selecting the change-point approach that appears to best estimate the expected trend prior to the diagnostic opportunity window, we then estimate the number of missed opportunities at each day during the window. Specifically, we estimate the expected trend using a simple linear function of time prior to diagnosis *t* by fitting the model *y*_*t*_ = *β*_0_ + *tβ*_1_ + *ε*_*t*_ over the interval [−*T, cp* − 1] and extrapolating *ŷ*_*t*_ over the interval [*cp*, −1]. We then compute the number of missed visits as the observed error *m*_*t*_ = *y*_*t*_ − *ŷ*_*t*_ using the observed count *y*_*t*_ over the interval [*cp*, −1].

#### Simulation models

We compare three different simulation approaches using two of the simulation algorithms described above. First, we use Algorithm 1 to draw missed visits that are independent at each period. Second, we use Algorithm 2 while setting *α* = 0. Third, we use Algorithm 2 while setting *α* = 1. Thus, our first simulation approach will describe the expected number of patients with a delay using uncorrelated draws, while the second and third models will roughly summarize the minimum and maximum number of patients potentially experiencing a delay, respectively using correlated draws. For simplicity, we refer to these approaches as *naïve* draws (algorithm 1), *minimal* patient-delays (algorithm 2, *α* = 0) and *maximal* patient-delays (algorithm 2, *α* = 1). Using these three simulation approaches, we estimate the following measures of the frequency of missed opportunities: the total number of visits representing a missed opportunity; the percent of missed opportunities occurring in inpatient, outpatient and emergency department settings; the percent of patients experiencing a missed opportunity; the mean number of missed opportunities each patient experienced; the mean duration of diagnostic delays (time from earliest missed opportunity to index diagnosis). We compute bootstrap-based 95% confidence intervals for each of these estimates by repeatedly redrawing 1,000 times which visits represented a missed opportunity, computing the above metrics, and using the 0.025 and 0.0975 percentile values.

#### Sensitivity analysis

It is possible that our SSD list and corresponding SSD visits may not fully capture all visits where a patient presented with symptoms of the disease. For example, there may be other SSD codes that are not directly specified in our SSD list. Alternatively, symptoms that occur during a visit may not be captured in the administrative discharge record because of recording errors (e.g., clinician fails to record symptom) or due to billing issues (e.g., no corresponding ICD code applied to the insurance claim). To address this potential limitation, we repeat our change-point analysis, obtain the estimates of the number of missed opportunities and employ our simulation models for all visits, instead of SSD visits only. This provides an upper bound on the number of potential missed opportunities, if SSD records are incomplete.

## Results

We identified 2,073 cases of tuberculosis, 359,625 cases of AMI, and 367,768 cases of stroke. Table 1 presents baseline characteristics for each of our study populations, including demographics, enrollment information and the number of observable visits per patient during the observation period prior to the index diagnosis. Figure 7 presents counts of SSD visits for each day leading up to the index diagnosis for each condition. There is a significant increase in SSD-related visits for all three conditions that occurs in the period before diagnosis.

**Table 1.** Baseline characteristics of the study cohorts for Stroke, AMI and Tuberculosis used for evaluating our simulation approach

|  | **Stroke** | **AMI** | **Tuberculosis** |
| --- | --- | --- | --- |
| **N** | 367,768 | 359,625 | 2,073 |
| **Age at Diagnosis (n (%))** |  |  |  |
| 18-30 | 17,972 (4.89%) | 4,174 (1.16%) | 252 (12.16%) |
| 31-45 | 61,820 (16.81%) | 45,835 (12.75%) | 620 (29.91%) |
| 46-55 | 121,995 (33.17%) | 131,604 (36.59%) | 588 (28.36%) |
| >55 | 165,981 (45.13%) | 178,012 (49.50%) | 613 (29.57%) |
| **Sex (n (%))** |  |  |  |
| Male | 186,966 (50.84%) | 249,899 (69.49%) | 988 (47.66%) |
| Female | 180,802 (49.16%) | 109,726 (30.51%) | 1,085 (52.34%) |
| **Enrollment Time Prior to Index (years)** |  |  |  |
| Mean | 3.62 | 3.70 | 3.86 |
| Median | 2.60 | 2.65 | 2.87 |
| Range | 0.49 - 17.01 | 0.49 - 17.01 | 1.00 - 15.79 |
| Count ≥ 1 year (n (%)) | 306,800 (83.42%) | 302,026 (83.98%) | 2,073 (100.00%) |
| Count ≥ 1.5 years (n (%)) | 260,581 (70.85%) | 257,661 (71.65%) | 1,719 (82.92%) |
| Count ≥ 2 years (n (%)) | 221,360 (60.19%) | 219,651 (61.08%) | 1,419 (68.45%) |
| Count ≥ 3 years (n (%)) | 162,047 (44.06%) | 161,224 (44.83%) | 997 (48.09%) |
| **Number of Visits in Period Prior to Diagnosis (i.e. 365 days prior for tuberculosis and 180 days prior for stroke and AMI) (n (%))** |  |  |  |
| 0 | 52,737 (14.34%) | 63,141 (17.56%) | 52 (2.51%) |
| 1-5 | 124,630 (33.89%) | 137,677 (38.28%) | 251 (12.11%) |
| 6-10 | 71,075 (19.33%) | 66,292 (18.43%) | 345 (16.64%) |
| 11-20 | 64,189 (17.45%) | 52,606 (14.63%) | 606 (29.23%) |
| > 20 | 55,137 (14.99%) | 39,909 (11.10%) | 819 (39.51%) |

**Figure 7.**
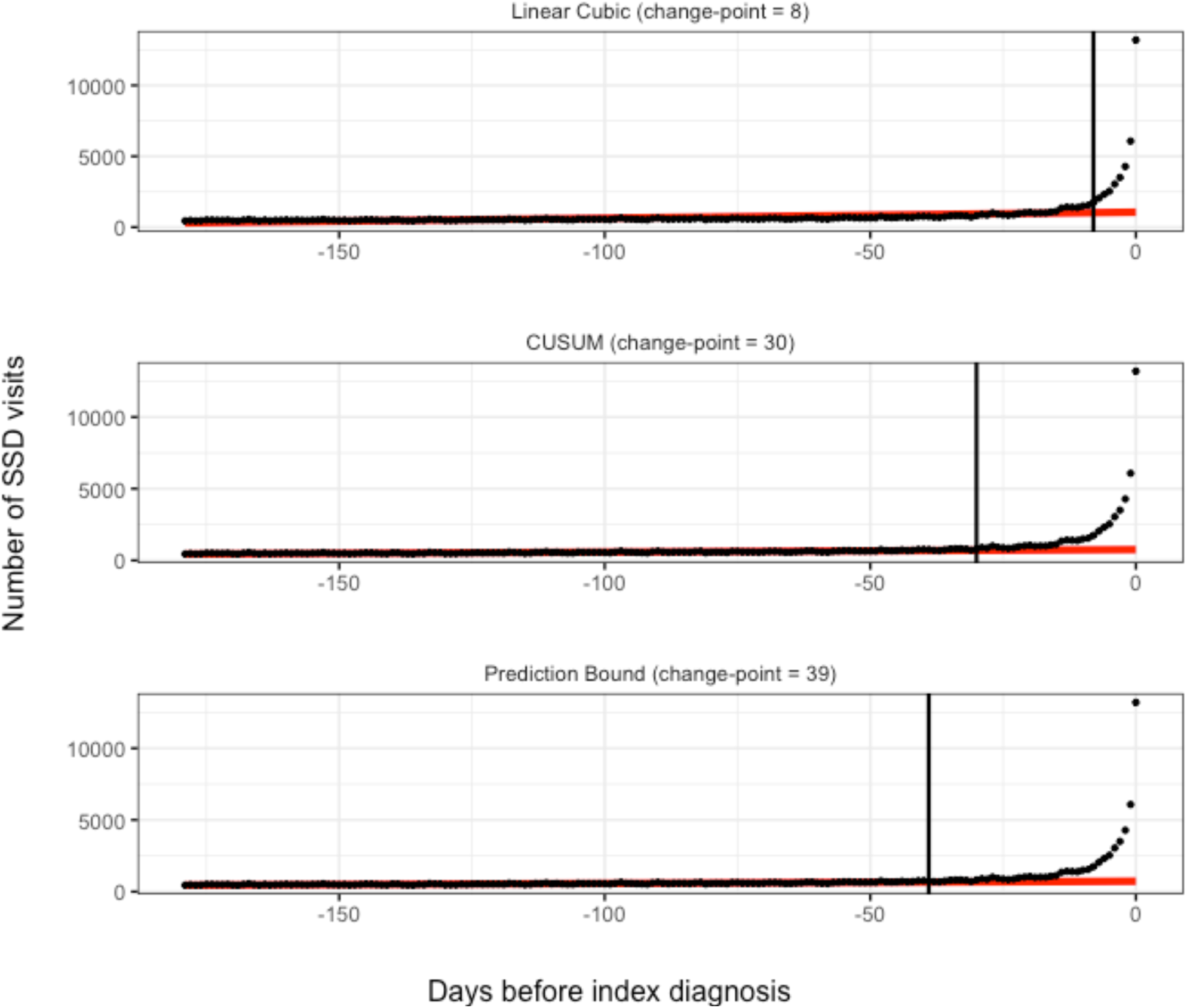
Counts of SSD visits each day prior to the index diagnosis. For each disease of interest there is an upward spike in the occurrence of healthcare visits with SSDs in the period just preceding the index diagnosis. The black vertical line represents the estimated change-point separating the diagnostic opportunity window from the prior crossover period. The red line represents the expected level of healthcare utilization (i.e., estimated to occur in absence of diagnostic delays).

For each condition, we estimated the change-point, expected number of baseline visits, and the number of missed visits using 3 different change-point detection approaches. Supplementary Table 3 and Supplementary Figures 1-3 present the resulting change-points, summaries of the number of missed opportunities and a visualization of the diagnostic opportunity window and expected number of SSD visits. For each condition, we identified the change-point approach that appeared to best fit the pattern of SSD visits based on the following criteria: (1) minimal MSE before the change-point, (2) mean error right before the change-point that is near zero. Supplementary Table 3 also lists these evaluation criteria for each of the methods and the three diseases considered. Note: each of these change-points that were “learned” by our change-point fitting approach are consistent with time periods that have been used to identify missed opportunities associated with each of these diseases.

For tuberculosis we selected the linear-cubic model which identified the change-point at 113 days prior to diagnosis. For AMI we selected the prediction bound model, which resulted in a change-point at 39 days before diagnosis. For stroke we also selected the prediction bound model, which resulted in a change-point at 38 days before diagnosis. Table 2 summarizes these results for each condition and the final selected change-point approach. Figure 7 also provides visualizations of the SSD visit counts along with the expected trend and corresponding opportunity window. Supplementary Table 4 provides the final estimates of the number of missed opportunities at each time period that was used in the simulation models.

**Table 2.** Selected Simulation Results. Estimates of the frequency of missed opportunities and duration of delays using different simulation algorithms

|  | Stroke | AMI | Tuberculosis |
| --- | --- | --- | --- |
| **Change Point (Start of diagnostic opportunity Window)** | 39 | 40 | 114 |
| **Total Number of Missed Opportunities During Delay Window (% of SSD visits during delay window)** | 41,577 (60.55%) | 103,874 (40.93%) | 6,444 (58.15%) |
| **Percent of Missed Opportunities in Outpatient Settings** |  |  |  |
| Naïve Approach | 62.64 (62.42 - 62.88) | 75.83 (75.68 - 75.99) | 89.23 (88.83 - 89.66) |
| Minimal Delays Approach | 63.04 (62.80 - 63.30) | 76.41 (76.25 - 76.56) | 90.25 (89.82 - 90.72) |
| Maximal Delays Approach | 61.52 (61.34 - 61.71) | 74.57 (74.44 - 74.70) | 88.58 (88.21 - 88.97) |
| **Percent of Missed Opportunities in Inpatient Settings** |  |  |  |
| Naïve Approach | 4.37 (4.25 - 4.48) | 5.89 (5.79 - 5.98) | 5.03 (4.72 - 5.31) |
| Minimal Delays Approach | 4.26 (4.15 - 4.37) | 5.86 (5.77 - 5.95) | 4.89 (4.61 - 5.17) |
| Maximal Delays Approach | 4.50 (4.40 - 4.61) | 5.90 (5.81 - 5.98) | 5.07 (4.81 - 5.32) |
| **Percent of Missed Opportunities in ED Settings** |  |  |  |
| Naïve Approach | 33.00 (32.78 - 33.21) | 18.28 (18.15 - 18.41) | 5.74 (5.40 - 6.05) |
| Minimal Delays Approach | 32.70 (32.47 - 32.92) | 17.73 (17.60 - 17.87) | 4.86 (4.52 - 5.18) |
| Maximal Delays Approach | 33.98 (33.80 - 34.16) | 19.53 (19.42 - 19.64) | 6.35 (6.07 - 6.60) |
| **Percent of Patients Experiencing Missed Opportunities (95% CI)** |  |  |  |
| Naïve Approach | 7.46 (7.43 - 7.48) | 18.50 (18.45 - 18.54) | 78.11 (77.33 - 78.87) |
| Minimal Delays Approach | 6.93 (6.91 - 6.96) | 16.01 (15.96 - 16.07) | 63.90 (62.81 - 65.03) |
| Maximal Delays Approach | 8.29 (8.28 - 8.31) | 21.31 (21.28 - 21.34) | 82.28 (81.96 - 82.59) |
| **Mean Number of Missed Opportunities among Patients Missed (95% CI)** |  |  |  |
| Naïve Approach | 1.52 (1.51 - 1.52) | 1.56 (1.56 - 1.57) | 3.98 (3.94 - 4.02) |
| Minimal Delays Approach | 1.63 (1.62 - 1.64) | 1.80 (1.80 - 1.81) | 4.87 (4.78 - 4.95) |
| Maximal Delays Approach | 1.36 (1.36 - 1.37) | 1.36 (1.35 - 1.36) | 3.78 (3.76 - 3.79) |
| **Mean Duration (days) of Missed Opportunities among Patients Missed (95% CI)** |  |  |  |
| Naïve Approach | 7.41 (7.38 - 7.44) | 8.10 (8.08 - 8.12) | 39.86 (39.31 - 40.37) |
| Minimal Delays Approach | 6.72 (6.67 - 6.76) | 6.69 (6.64 - 6.73) | 34.35 (33.52 - 35.15) |
| Maximal Delays Approach | 7.64 (7.63 - 7.66) | 8.21 (8.20 - 8.22) | 44.47 (44.16 - 44.75) |

For each condition we applied three different simulation approaches (naïve, minimal, and maximal), as described above, to identify which visits represented a missed opportunity. Table 2 presents the results of these simulations in terms of the settings where missed opportunities occurred (outpatient, inpatient or ED), the percentage of patients experiencing at least one missed opportunity, the mean number of missed opportunities such patients experienced and the average duration of diagnostic delays. The results shown in this table demonstrate the sensitivity of these estimates to the type of simulation algorithm applied.

In general, across all three diseases the majority of diagnostic opportunities appeared to occur in outpatient settings, representing around 90% of missed opportunities for tuberculosis, 75% for AMI and 60% for stroke. The ED was the second most common setting for misses representing around 30% for stroke, 20% for AMI and 5% for tuberculosis. For all three diseases, around 4-6% of missed opportunities occurred in inpatient settings. The type of simulation algorithm did not dramatically alter these estimates, with the percentage by type of setting only differing by a percentage point or two across algorithms for each disease. However, the maximal patient-delays approach did tend to result in fewer missed opportunities ascribed to outpatient settings, while the minimal approach resulted in slightly more and the naïve approach tended to be in between algorithms 2 and 3.

The differences between the algorithms are reflected more clearly in the estimates of the percentage of patients experiencing a missed opportunity, the number of missed opportunities ascribed to each patient who was missed, and the duration of diagnostic delays among patients who were delayed. The maximal tended to result in the largest number of patients experiencing a miss, as expected; the minimal resulted in the fewest while the naïve approach was somewhere in between. Patients with tuberculosis were most likely to experience a missed opportunity, with between 63.9% - 82.3% of patients experiencing at least one missed opportunity, depending on the algorithm, while patients with stroke were least likely, with 6.9% - 8.3% of patients experiencing a missed opportunity across the different algorithms. These differences across algorithms are consistent with the intuition described above where the maximal approach essentially identifies more patients (i.e., patients not previously drawn) at each successive draw, while the minimal approach tries to identify fewer patients (i.e., previously drawn patients).

Similarly, the correlated nature of successive draws results in differences in the average number of missed opportunities those patients who experience a delayed diagnosis were assigned. The minimal approach, which identified the fewest number of patients with a missed opportunity, resulted in more missed opportunities assigned to each patient who did experience at least one missed opportunity, compared to the maximal approach, which tended to identify the greatest number of individuals with at least one missed opportunity. Patients with tuberculosis experienced the greatest number of missed opportunities, with between 3.78 missed opportunities per patient for the maximal approach versus 4.87 missed opportunities for the minimal approach. Patients with AMI and stroke who experienced a missed opportunity had a similar number of missed opportunities across algorithms (1.36-1.63 per patient for stroke and 1.36-1.80 for AMI).

A somewhat counterintuitive result occurs across algorithms related to the estimated duration of diagnostic delays. The minimal approach, which identifies the fewest number of total patients with a diagnostic delay and results in more missed opportunities per patient, tended to result in a shorter average duration of diagnostic delays, compared to the maximal approach which produced longer diagnostic delays on average. This result is due to the skewed nature of SSD visits (see Figure 7) and the correlated nature of subsequent draws between the algorithms. Because the minimal approach attempts to draw missed opportunities at each subsequent time period from patients who have already been drawn (i.e., a longer delay), there is a clustering of the earlier SSD visits among patients with already long duration of delays; those visits which occur further before the index date are more likely to be assigned to patients already having a longer delay, while newly drawn patients are more likely to be selected corresponding to visits closer to the index date. Thus, the minimal approach generates a relatively small number of patients who have long delays but with many visits further before the index date while producing relatively more patients with short delays closer to the index. This difference between algorithms has a relatively large impact on the average duration for diagnostic delays in the case of tuberculosis, with the average duration of 34.35 days for the minimal approach versus 44.47 days for the maximal approach. However, the difference between algorithms is less dramatic for stroke (6.72 vs 7.64) and AMI (6.69 vs 8.21).

We also conducted a sensitivity analysis of the SSD list used to identify potential missed opportunities. In particular, we repeated our change-point analysis, estimated the observed and expected visit trends, computed the estimated numbers of missed opportunities and repeated all simulation analysis using all visits, instead of SSD visits only. These results are presented in Supplementary Table 10. In general, this analysis resulted in a much greater estimated number of missed opportunities (more than 5 times as many for stroke, 1.63 times for AMI and 1.52 times for tuberculosis). The general trends across algorithms, described above, was the same with 3 notable exceptions. First, a greater proportion of missed opportunities were estimated to occur in outpatient settings. Second, a greater percentage of patients were estimated to have experienced a missed opportunity; this result was most exaggerated for AMI and stroke. Third, the mean number of missed opportunities and duration of delays among those patients who experienced a delay was not consistently different; in some cases, the number of delays per patient and duration of delays increased, while in others it was the same or even decreased. Thus, this type of sensitivity analysis may help to broaden the number of missed opportunities and patients with missed opportunities identified, but may not dramatically change the dynamics of missed opportunities among patients identified to have a delay in terms of the number of missed opportunities they are estimated to experience or the estimated duration of delay.

## Discussion

In this paper, we presented a general simulation-based approach to estimate individual-level measures of missed diagnostic opportunities from longitudinal health records. Specifically, this approach allows one to estimate the frequency of missed opportunities at an aggregate level, along with individual-level metrics such as the number of patients experiencing a missed opportunity, the settings where missed opportunities occur, the number of missed opportunities that individual patients experience, and the duration of diagnostic delays. We applied these methods to TB, AMI, and stroke, and consistent with prior investigations for these diseases, we identified a significant number of missed opportunities associated with these diseases. We also demonstrated that a range of results may be generated based on different evaluation criteria.

The simulation approach we describe, unlike many prior approaches to study diagnostic delays, such as retrospective chart reviews, autopsy studies, or malpractice claims is less costly and time consuming and can be applied to virtually any disease captured by longitudinal patient records. Moreover, our approach provides a high degree of flexibility in terms of estimation procedures, algorithms for selecting missed opportunities and output measures. While other approaches have used similar longitudinal data sources and a type of “look-back” approach to study diagnostic delays, these approaches often have a number of methodological limitations that our approach is designed to address.^8,9,11,13,16^ First, these studies are often unable to compute individual-level patient metrics, such as delay duration or frequency of misses in individual patients, as these metrics typically require analyzing individual patient records. Second, these studies generally require expert specification of criteria to define a delay (e.g., time prior to diagnosis). Third, the approaches very often do not account for the fact that many signs and symptoms occurring before diagnosis may be unrelated to the disease of interest and can be expected to occur even in absence of diagnostic delays; thus, prior approaches may not distinguish between likely missed opportunities and coincidental visits. To our knowledge, only a few prior studies have attempted to control for this coincidental level of care^17-19^ and our approach can be viewed as an extension of these prior methods.

We presented three simulation algorithms and applied two of these to the diseases of interest. Given our findings, which demonstrated differing results across algorithms, we offer the following guidance to future investigators wishing to use this approach. First, because the application of these methods and data sources to study diagnostic delays is relatively novel, alternative algorithms may be used as means to provide a sensitivity analysis around empirical estimates. In cases where relatively little is known about the correlation between patient revisits and the likelihood of an individual visit representing a delay, we recommend the general approach utilized in this paper. Namely, the naïve approach (Algorithm 1) should be considered as the baseline or default estimate. This algorithm places the fewest assumptions on the simulation process, and is conceptually the most straightforward. However, investigators may also consider Algorithm 2, while setting *α* = 0 and *α* = 1 to provide bounds related to algorithmic sensitivity. Second, in situations where more information is known about the correlation between repeated missed opportunities prior to diagnosis (e.g., symptoms are known to persist among patients who are delayed), expert evaluation may be used to determine if α, in Algorithm 2, should be set closer to 0 or 1. Finally, as future investigations make use of these approaches, or as validation studies are conducted, such information may be used to develop a more realistic specification for the generalized algorithm (Algorithm 3) that may allow it to be better customized to disease-specific contexts.

One of the most important contributions of our approach is that our method explicitly attempts to account for the expected healthcare that may coincidently occur prior to the index diagnosis of a disease of interest, while also providing a means for generating individual-level delay analysis. For example, not all respiratory events that occur prior to a tuberculosis diagnosis may be a direct result of tuberculosis, and many visits that appear to be potential missed opportunities may be coincidental. Failure to account for these expected trends may lead to significant overestimates in the frequency of diagnostic opportunities. However, prior investigations that have used similar data sources and approaches to identify individual missed diagnostic opportunities have typically labeled all events that meet pre-specified criteria (e.g., dizziness before stroke) as a “missed opportunity.” Consequently, these approaches, are often paired with additional criteria (e.g., treat and release ED visits^8,9,11^) to ensure greater specificity but come at the cost of decreasing the sensitivity in identifying missed opportunities. Attempts have been made to account for observed patterns of symptomatic visits relative to what would have been expected using either other visits^11^ or using a crossover period prior to when delays may be expected to occur,^17^ as we have proposed. However, these approaches have still been unable to compute individual-level-patient metrics, such as delay duration or frequency of misses in individual patients.

Another primary advantage of our approach is that it provides a fairly flexible set of criteria for guiding the estimation process of diagnostic delays. First, numerous estimation procedures can be used to estimate the simulation parameters described above, including the change-point for the diagnostic opportunity window and the trend in expected SSD visits. Second, we have presented three basic algorithms for drawing/simulating missed opportunities. Our generalized algorithm presents a customizable weighting parameter that can be used to adapt the simulation to a particular disease of interest. Multiple other extensions are possible, and the algorithms here can be customized for more complex scenarios. For example, a sequential selection criterion such as – *a patient who experiences SSD A then SSD B is more likely to represent a diagnostic delay than SSD B before SSD A*. Thus, our results present a simulation outline upon which future investigations can build.

Results from our different simulation approaches demonstrate that a range of estimates may be generated based on how one chooses to define the correlation structure between missed opportunities identified across sequential draws within the simulation. For example, the type of algorithm selected resulted in differences in the percent of individuals identified to have a delay, the setting where missed opportunities occurred (inpatient, outpatient, ED), the duration of delay and number of missed opportunities per patient. In some cases (e.g., mean duration of delays with tuberculosis), the difference in estimates between algorithms can be quite large. Thus, clinical knowledge should still be employed to guide the estimation process and identify the simulation approaches that best suit the particular disease. However, the simulations we present can provide bounds on the range of plausible results at an individual level. In addition, our sensitivity analysis using all visits prior to diagnosis may be one approach to provide an upper bound on the estimated number of missed opportunities.

A final benefit of our simulation approach is that it may provide a unified approach for quantifying and comparing the frequency and duration of diagnostic delays across a variety of diseases in a more reproducible fashion. As noted above, there are considerable challenges when attempting to compare estimates of diagnostic delays across studies where differing methods and study populations are used. Our approach may provide one way to generate comparable results across diseases and studies, allowing investigators to directly compare which diseases may have a longer or shorter delay process and/or a greater/lesser frequency of delays. For example, in our applications, missed opportunities were far more common for tuberculosis compared to stroke and even less common for AMI. Average delays for AMI and stroke were similar at around 7-8 days versus tuberculosis, which was around 40 days. For all three diseases, only around 5% of the missed opportunities we identified occurred in inpatient settings, but there were considerable differences between ED and other outpatient care. These and similar metrics may also be useful for benchmarking purposes or providing a measure of *diagnostic efficiency* (e.g., how many healthcare resources are typically required to make a correct diagnosis) across diseases. Such measures may be useful for policy makers wishing to evaluate the relative importance of delays across a wide range of diseases using widely available data sources.

## Limitations

There are a number of limitations with the simulation approach we present here. First, our approach generally requires a large data source of longitudinal patient records, especially for diseases that are relatively rare. While such records are often readily available in the form of administrative claims, discharge records or other observational data sources, such data typically do not contain the types of granular information necessary for in-depth validation of delays (e.g., clinic notes or vital signs). A second limitation of our approach is that the missed opportunities identified do not necessarily imply diagnostic delays have occurred, and even “likely” missed opportunities for diagnosis may be unavoidable even in settings of ideal patient care. Our approach is simply designed to detect missed opportunities based on “excess” SSD visits that deviate from expectations to a statistically meaningful degree. However, we cannot assume that a healthcare provider would reasonably be expected to diagnose each of these cases, and our approach does not incorporate harms that may have resulted. Similarly, our approach may miss longer delays or those that do not generate a significant signal in aggregate visit counts. For example, the addition of a handful of missed opportunities occurring outside the delay opportunity window are unlikely to impact the detection of the optimal change-point. Thus, there may exist some patients whose delay is not completely captured by our approach. Finally, the approach outlined above assumes the defined SSD set is relatively complete. If a significant number of SSDs are unknown or not included in the primary analysis, results may be significantly underestimated. However, we also presented a sensitivity analysis using all visits that may be used to compute upper bounds on the number of delays and provide guidance on the relative completeness of the defined SSD set.

## Conclusions

The simulation approach presented here provides an intuitive, flexible and broadly applicable framework that can be used to identify missed opportunities and study diagnostic delays using large longitudinal data sources. This approach is less costly and time intensive than traditional methods to study diagnostic delays. It builds upon recent efforts to utilize large real-world datasets to study diagnostic delays, but also addresses many of the limitations present in prior study designs. Our results demonstrate consistency with prior investigations of diagnostic delays, but also provide a means to generate future results for different diseases and study populations. Moreover, we outlined a number of flexible extensions upon which future investigations and clinical expertise may be used to expand and refine our general approach to individual diseases.

## Data Availability

This study used data from the IBM MarketScan Research Databases. These are commercially available de-itentified administrative claims datasets, that can be obtained from IBM MarketScan. Other researchers can purchase access to these data through IBM MarketScan (https://www.ibm.com/products/marketscan-research-databases). The collection of these data along with the process for de-identifying records is the intellectual property of IBM MarketScan. The study authors have obtained a license to use these data, and this research was conducted in accordance with the corresponding Data Use Agreement.

