## Supplemental Material for "A comprehensive framework to estimate the frequency, duration and risk factors for diagnostic delays using simulation-based methods"

**SUPPLEMENTARY MATERIAL**

**Supplementary Table 1. Disease-specific index diagnosis codes**

| **Disease** | **ICD9-CM** | **ICD10-CM** |
| --- | --- | --- |
| **Stroke** | 430, 431, 432, 432.0, 432.1, 432.9, 433.01, 433.11, 433.21, 433.31, 433.81, 433.91, 434, 434.0, 434.00, 434.01, 434.1, 434.10, 434.11, 434.9, 434.90, 434.91, 346.60, 346.61, 346.62, 346.63, 436, 435, 435.0, 435.1, 435.2, 435.3, 435.8, 435.9 | G43.609, G43.619, G43.601, G43.611, I60.9, I61.9, I62.1, I62.00, I62.9, I63.22, I63.139, I63.239, I63.019, I63.119, I63.219, I63.59, I63.20, I66.09, I66.19, I66.29, I63.30, I66.9, I63.40, I63.50, G45.0, G45.8, G45.1, G45.9, I67.848, I67.89, G45.2, G46.0, G46.1, G46.2, I60.00, I60.01, I60.02, I60.10, I60.11, I60.12, I60.20, I60.21, I60.22, I60.30, I60.31, I60.32, I60.4, I60.50, I60.51, I60.52, I60.6, I60.7, I60.8, I61.0, I61.1, I61.2, I61.3, I61.4, I61.5, I61.6, I61.8, I62.01, I62.02, I62.03, I63.00, I63.011, I63.012, I63.02, I63.031, I63.032, I63.039, I63.09, I63.10, I63.111, I63.112, I63.12, I63.131, I63.132, I63.19, I63.211, I63.212, I63.231, I63.232, I63.29, I63.311, I63.312, I63.319, I63.321, I63.322, I63.329, I63.331, I63.332, I63.339, I63.341, I63.342, I63.349, I63.39, I63.411, I63.412, I63.419, I63.421, I63.422, I63.429, I63.431, I63.432, I63.439, I63.441, I63.442, I63.449, I63.49, I63.511, I63.512, I63.519, I63.521, I63.522, I63.529, I63.531, I63.532, I63.539, I63.541, I63.542, I63.549, I63.6, I63.8, I63.9, I66.01, I66.02, I66.03, I66.11, I66.12, I66.13, I66.21, I66.22, I66.23, I66.3, I66.8, I67.841 |
| **AMI** | 410.0, 410.00, 410.01, 410.02, 410.1, 410.10, 410.11, 410.12, 410.2, 410.20, 410.21, 410.22, 410.3, 410.30, 410.31, 410.32, 410.4, 410.40, 410.41, 410.42, 410.5, 410.50, 410.51, 410.52, 410.6, 410.60, 410.61, 410.62, 410.7, 410.70, 410.71, 410.72, 410.8, 410.80, 410.81, 410.82, 410.9, 410.90, 410.91, 410.92 | I21.01, I21.02, I21.09, I21.11, I21.19, I21.21, I21.29, I21.3, I21.4, I21.9, I21.A1, I21.A9, I22.0, I22.1, I22.2, I22.8, I22.9 |
| **Tuberculosis** | 010, 010.0, 010.00, 010.01, 010.02, 010.03, 010.04, 010.05, 010.06, 010.1, 010.10, 010.11, 010.12, 010.13, 010.14, 010.15, 010.16, 010.8, 010.80, 010.81, 010.82, 010.83, 010.84, 010.85, 010.86, 010.9, 010.90, 010.91, 010.92, 010.93, 010.94, 010.95, 010.96, 011, 011.0, 011.00, 011.01, 011.02, 011.03, 011.04, 011.05, 011.06, 011.1, 011.10, 011.11, 011.12, 011.13, 011.14, 011.15, 011.16, 011.2, 011.20, 011.21, 011.22, 011.23, 011.24, 011.25, 011.26, 011.3, 011.30, 011.31, 011.32, 011.33, 011.34, 011.35, 011.36, 011.4, 011.40, 011.41, 011.42, 011.43, 011.44, 011.45, 011.46, 011.5, 011.50, 011.51, 011.52, 011.53, 011.54, 011.55, 011.56, 011.6, 011.60, 011.61, 011.62, 011.63, 011.64, 011.65, 011.66, 011.7, 011.70, 011.71, 011.72, 011.73, 011.74, 011.75, 011.76, 011.8, 011.80, 011.81, 011.82, 011.83, 011.84, 011.85, 011.86, 011.9, 011.90, 011.91, 011.92, 011.93, 011.94, 011.95, 011.96, 012, 012.0, 012.00, 012.01, 012.02, 012.03, 012.04, 012.05, 012.06, 012.1, 012.10, 012.11, 012.12, 012.13, 012.14, 012.15, 012.16, 012.2, 012.20, 012.21, 012.22, 012.23, 012.24, 012.25, 012.26, 012.3, 012.30, 012.31, 012.32, 012.33, 012.34, 012.35, 012.36, 012.8, 012.80, 012.81, 012.82, 012.83, 012.84, 012.85, 012.86, 018, 018.0, 018.00, 018.01, 018.02, 018.03, 018.04, 018.05, 018.06, 018.8, 018.80, 018.81, 018.82, 018.83, 018.84, 018.85, 018.86, 018.9, 018.90, 018.91, 018.92, 018.93, 018.94, 018.95, 018.96 | A15, A15.0, A15.4, A15.5, A15.6, A15.7, A15.8, A15.9, A19, A19.0, A19.1, A19.2, A19.8, A19.9 |

**Supplementary Table 2. Disease-specific SSD diagnosis codes**

| **Disease** | **SSD** | **ICD-9-CM** | **ICD-10-CM** |
| --- | --- | --- | --- |
| **Stroke** | Headache | 339.00, 339.01, 339.02, 339.03, 339.04, 346.0, 346.00, 346.01, 346.02, 346.03, 346.1, 346.10, 346.11, 346.12, 346.13, 346.2, 346.20, 346.21, 346.22, 346.23, 346.30, 346.31, 346.32, 346.33, 346.40, 346.41, 346.42, 346.43, 346.50, 346.51, 346.52, 346.53, 346.70, 346.71, 346.72, 346.73, 346.8, 346.80, 346.81, 346.82, 346.83, 346.9, 346.90, 346.91, 346.92, 346.93, 339.05, 339.09, 339.10, 339.11, 339.12, 339.20, 339.21, 339.22, 339.3, 339.41, 339.42, 339.43, 339.44, 339.81, 339.82, 339.83, 339.84, 339.85, 339.89, 784.0 | G44.009, G44.019, G44.029, G44.039, G44.049, G44.059, G44.099, G44.209, G44.219, G44.221, G44.229, G44.309, G44.319, G44.329, G44.41, G44.51, G44.52, G44.53, G44.59, G44.81, G44.82, G44.83, G44.84, G44.85, G44.89, G43.109, G43.119, G43.101, G43.111, G43.009, G43.019, G43.001, G43.011, G43.809, G43.A0, G43.B0, G43.C0, G43.D0, G43.819, G43.A1, G43.B1, G43.C1, G43.D1, G43.801, G43.811, G43.409, G43.419, G43.401, G43.411, G43.829, G43.839, G43.821, G43.831, G43.509, G43.519, G43.501, G43.511, G43.709, G43.719, G43.701, G43.711, G43.809, G43.819, G43.801, G43.811, G43.909, G43.919, G43.901, G43.911, G44.1, R51 |
|  | Dizziness | 386.00, 386.01, 386.02, 386.03, 386.04, 386.10, 386.11, 386.12, 386.19, 386.2, 386.30, 386.31, 386.32, 386.33, 386.34, 386.35, 386.40, 386.41, 386.42, 386.43, 386.48, 386.50, 386.51, 386.52, 386.53, 386.54, 386.55, 386.56, 386.58, 386.8, 386.9, 780.4, 380.00, 380.01, 380.02, 380.03, 380.10, 380.11, 380.12, 380.13, 380.14, 380.15, 380.16, 380.21, 380.22, 380.23, 380.30, 380.31, 380.32, 380.39, 380.4, 380.50, 380.51, 380.52, 380.53, 380.81, 380.89, 380.9, 400, 384.01, 384.09, 384.1, 385.30, 385.31, 385.32, 385.33, 385.35, 385.82, 385.83, 385.89, 385.9, 388.00, 388.01, 388.02, 388.10, 388.11, 388.12, 388.2, 388.30, 388.31, 388.32, 388.40, 388.41, 388.42, 388.43, 388.44, 388.45, 388.5, 388.60, 388.61, 388.69, 388.70, 388.71, 388.72, 388.8, 388.9, 389.00, 389.01, 389.02, 389.03, 389.04, 389.05, 389.06, 389.08, 389.10, 389.11, 389.12, 389.13, 389.14, 389.15, 389.16, 389.17, 389.18, 389.2, 389.20, 389.21, 389.22, 389.7, 389.8, 389.9, V41.2, V41.3, V49.85, V53.2, V72.1, V72.11, V72.12, V72.19 | H61.009, H61.019, H61.029, H61.039, H60.00, H60.10, H60.319, H60.329, H60.399, H61.93, H60.339, H62.40, H60.20, H62.8X1, H60.399, H60.40, H60.509, H60.519, H60.529, H60.539, H60.549, H60.559, H60.599, H60.60, H60.8X1, H60.90, H61.109, H61.129, H61.119, H61.199, H61.23, H61.309, H61.399, H61.319, H61.399, H61.329, H61.819, H61.899, H61.93, H73.019, H73.099, H73.10, H71.93, H71.03, H71.13, H74.40, H71.23, H71.33, H74.8X9, H74.8X9, H74.8X9, H74.90, H81.09, H81.09, H81.09, H81.09, H81.09, H81.399, H81.13, H81.23, H81.319, H81.49, H83.09, H83.09, H83.09, H83.09, H83.09, H83.09, H83.19, H83.19, H83.19, H83.19, H83.19, H83.2X9, H83.2X9, H83.2X9, H83.2X9, H83.2X9, H83.2X9, H83.2X9, H83.2X9, H81.8X9, H82.9, H83.8X9, H81.93, H83.93, H93.099, H91.13, H93.019, H83.3X9, H91.8X9, H83.3X9, H91.23, H93.19, H93.19, H93.19, H93.249, H93.299, H93.229, H93.239, H93.299, H93.219, H93.299, H93.3X9, H92.10, G96.0, H92.20, H92.09, H92.09, H92.09, H93.8X9, H93.93, H90.2, H90.2, H90.2, H90.2, H90.2, H90.11, H90.12, H90.0, H90.2, H90.5, H90.3, H90.3, H90.41, H90.42, H90.5, H90.41, H90.42, H90.5, H90.41, H90.42, H90.3, H90.8, H90.71, H90.72, H90.6, H91.3, H91.8X9, H91.90, R42, Z97.4, H93.90, Z73.82, Z46.1, Z01.110, Z01.12, Z01.10, Z01.118 |
| **AMI** | Syncope | 780.2 | 795.5, 795.51, 795.52, 795.6, 795.79, V01.0, V01.1, V01.2, V01.3, V01.4, V01.5, V01.6, V01.7, V01.71, V01.79, V01.8, V01.81, V01.82, V01.83, V01.84, V01.89, V01.9, V02.0, V02.1, V02.2, V02.3, V02.4, V02.5, V02.51, V02.52, V02.53, V02.54, V02.59, V02.6, V02.60, V02.61, V02.62, V02.69, V02.7, V02.8, V02.9, V03.0, V03.1, V03.2, V03.3, V03.4, V03.5, V03.6, V03.7, V03.8, V03.81, V03.82, V03.89, V03.9, V04.0, V04.1, V04.2, V04.3, V04.4, V04.5, V04.6, V04.7, V04.8, V04.81, V04.82, V04.89, V05.0, V05.1, V05.2, V05.3, V05.4, V05.8, V05.9, V06.0, V06.1, V06.2, V06.3, V06.4, V06.5, V06.6, V06.8, V06.9, V28.6, V71.2, V71.82, V71.83, V73.0, V73.1, V73.2, V73.3, V73.4, V73.5, V73.6, V73.8, V73.81, V73.88, V73.89, V73.9, V73.98, V73.99, V74.0, V74.1, V74.2, V74.3, V74.4, V74.5, V74.6, V74.8, V74.9, V75.0, V75.1, V75.2, V75.3, V75.4, V75.5, V75.6, V75.7, V75.8, V75.9, 780.2 |
|  | Other Lower Respiratory Disease | 513.1, 514, 515, 516.0, 516.1, 516.2, 516.3, 516.30, 516.31, 516.32, 516.33, 516.34, 516.35, 516.36, 516.37, 516.4, 516.5, 516.61, 516.62, 516.63, 516.64, 516.69, 516.8, 516.9, 517.2, 517.8, 518.3, 518.4, 518.89, 519.4, 519.8, 519.9, 782.5, 786.00, 786.01, 786.02, 786.03, 786.04, 786.05, 786.06, 786.07, 786.09, 786.2, 786.3, 786.30, 786.31, 786.39, 786.4, 786.52, 786.6, 786.7, 786.8, 786.9, 793.1, 793.11, 793.19, 794.2, V12.6, V12.60, V12.61, V12.69, V42.6 | 795.5, 795.51, 795.52, 795.6, 795.79, V01.0, V01.1, V01.2, V01.3, V01.4, V01.5, V01.6, V01.7, V01.71, V01.79, V01.8, V01.81, V01.82, V01.83, V01.84, V01.89, V01.9, V02.0, V02.1, V02.2, V02.3, V02.4, V02.5, V02.51, V02.52, V02.53, V02.54, V02.59, V02.6, V02.60, V02.61, V02.62, V02.69, V02.7, V02.8, V02.9, V03.0, V03.1, V03.2, V03.3, V03.4, V03.5, V03.6, V03.7, V03.8, V03.81, V03.82, V03.89, V03.9, V04.0, V04.1, V04.2, V04.3, V04.4, V04.5, V04.6, V04.7, V04.8, V04.81, V04.82, V04.89, V05.0, V05.1, V05.2, V05.3, V05.4, V05.8, V05.9, V06.0, V06.1, V06.2, V06.3, V06.4, V06.5, V06.6, V06.8, V06.9, V28.6, V71.2, V71.82, V71.83, V73.0, V73.1, V73.2, V73.3, V73.4, V73.5, V73.6, V73.8, V73.81, V73.88, V73.89, V73.9, V73.98, V73.99, V74.0, V74.1, V74.2, V74.3, V74.4, V74.5, V74.6, V74.8, V74.9, V75.0, V75.1, V75.2, V75.3, V75.4, V75.5, V75.6, V75.7, V75.8, V75.9, 513.1, 514, 515, 516.0, 516.1, 516.2, 516.3, 516.30, 516.31, 516.32, 516.33, 516.34, 516.35, 516.36, 516.37, 516.4, 516.5, 516.61, 516.62, 516.63, 516.64, 516.69, 516.8, 516.9, 517.2, 517.8, 518.3, 518.4, 518.89, 519.4, 519.8, 519.9, 782.5, 786.00, 786.01, 786.02, 786.03, 786.04, 786.05, 786.06, 786.07, 786.09, 786.2, 786.3, 786.30, 786.31, 786.39, 786.4, 786.52, 786.6, 786.7, 786.8, 786.9, 793.1, 793.11, 793.19, 794.2, V12.6, V12.60, V12.61, V12.69, V42.6 |
|  | Other Gastrointestinal Disorders | 538, 558.1, 558.2, 564.0, 564.00, 564.01, 564.02, 564.09, 564.1, 564.5, 564.7, 564.8, 564.81, 564.89, 564.9, 568.0, 568.81, 568.82, 568.89, 568.9, 569.81, 569.82, 569.83, 569.84, 569.85, 569.86, 569.87, 569.89, 569.9, 579.0, 579.1, 579.2, 579.8, 579.9, 787.1, 787.2, 787.20, 787.21, 787.22, 787.23, 787.24, 787.29, 787.3, 787.4, 787.5, 787.6, 787.60, 787.61, 787.62, 787.63, 787.7, 787.9, 787.91, 787.99, 789.2, 789.3, 789.30, 789.31, 789.32, 789.33, 789.34, 789.35, 789.36, 789.37, 789.39, 789.4, 789.40, 789.41, 789.42, 789.43, 789.44, 789.45, 789.46, 789.47, 789.49, 789.9, 792.1, 793.4, 793.6, V12.7, V12.70, V12.79, V41.6, V44.1, V44.2, V44.3, V44.4, V45.3, V47.3, V53.5, V53.50, V53.51, V53.59, V55.1, V55.2, V55.3, V55.4 | 795.5, 795.51, 795.52, 795.6, 795.79, V01.0, V01.1, V01.2, V01.3, V01.4, V01.5, V01.6, V01.7, V01.71, V01.79, V01.8, V01.81, V01.82, V01.83, V01.84, V01.89, V01.9, V02.0, V02.1, V02.2, V02.3, V02.4, V02.5, V02.51, V02.52, V02.53, V02.54, V02.59, V02.6, V02.60, V02.61, V02.62, V02.69, V02.7, V02.8, V02.9, V03.0, V03.1, V03.2, V03.3, V03.4, V03.5, V03.6, V03.7, V03.8, V03.81, V03.82, V03.89, V03.9, V04.0, V04.1, V04.2, V04.3, V04.4, V04.5, V04.6, V04.7, V04.8, V04.81, V04.82, V04.89, V05.0, V05.1, V05.2, V05.3, V05.4, V05.8, V05.9, V06.0, V06.1, V06.2, V06.3, V06.4, V06.5, V06.6, V06.8, V06.9, V28.6, V71.2, V71.82, V71.83, V73.0, V73.1, V73.2, V73.3, V73.4, V73.5, V73.6, V73.8, V73.81, V73.88, V73.89, V73.9, V73.98, V73.99, V74.0, V74.1, V74.2, V74.3, V74.4, V74.5, V74.6, V74.8, V74.9, V75.0, V75.1, V75.2, V75.3, V75.4, V75.5, V75.6, V75.7, V75.8, V75.9, 538, 558.1, 558.2, 564.0, 564.00, 564.01, 564.02, 564.09, 564.1, 564.5, 564.7, 564.8, 564.81, 564.89, 564.9, 568.0, 568.81, 568.82, 568.89, 568.9, 569.81, 569.82, 569.83, 569.84, 569.85, 569.86, 569.87, 569.89, 569.9, 579.0, 579.1, 579.2, 579.8, 579.9, 787.1, 787.2, 787.20, 787.21, 787.22, 787.23, 787.24, 787.29, 787.3, 787.4, 787.5, 787.6, 787.60, 787.61, 787.62, 787.63, 787.7, 787.9, 787.91, 787.99, 789.2, 789.3, 789.30, 789.31, 789.32, 789.33, 789.34, 789.35, 789.36, 789.37, 789.39, 789.4, 789.40, 789.41, 789.42, 789.43, 789.44, 789.45, 789.46, 789.47, 789.49, 789.9, 792.1, 793.4, 793.6, V12.7, V12.70, V12.79, V41.6, V44.1, V44.2, V44.3, V44.4, V45.3, V47.3, V53.5, V53.50, V53.51, V53.59, V55.1, V55.2, V55.3, V55.4 |
|  | Nonspecific Chest Pain | 786.50, 786.51, 786.59 | 795.5, 795.51, 795.52, 795.6, 795.79, V01.0, V01.1, V01.2, V01.3, V01.4, V01.5, V01.6, V01.7, V01.71, V01.79, V01.8, V01.81, V01.82, V01.83, V01.84, V01.89, V01.9, V02.0, V02.1, V02.2, V02.3, V02.4, V02.5, V02.51, V02.52, V02.53, V02.54, V02.59, V02.6, V02.60, V02.61, V02.62, V02.69, V02.7, V02.8, V02.9, V03.0, V03.1, V03.2, V03.3, V03.4, V03.5, V03.6, V03.7, V03.8, V03.81, V03.82, V03.89, V03.9, V04.0, V04.1, V04.2, V04.3, V04.4, V04.5, V04.6, V04.7, V04.8, V04.81, V04.82, V04.89, V05.0, V05.1, V05.2, V05.3, V05.4, V05.8, V05.9, V06.0, V06.1, V06.2, V06.3, V06.4, V06.5, V06.6, V06.8, V06.9, V28.6, V71.2, V71.82, V71.83, V73.0, V73.1, V73.2, V73.3, V73.4, V73.5, V73.6, V73.8, V73.81, V73.88, V73.89, V73.9, V73.98, V73.99, V74.0, V74.1, V74.2, V74.3, V74.4, V74.5, V74.6, V74.8, V74.9, V75.0, V75.1, V75.2, V75.3, V75.4, V75.5, V75.6, V75.7, V75.8, V75.9, 786.50, 786.51, 786.59 |
|  | Malaise and Fatigue | 780.7, 780.71, 780.79 | 795.5, 795.51, 795.52, 795.6, 795.79, V01.0, V01.1, V01.2, V01.3, V01.4, V01.5, V01.6, V01.7, V01.71, V01.79, V01.8, V01.81, V01.82, V01.83, V01.84, V01.89, V01.9, V02.0, V02.1, V02.2, V02.3, V02.4, V02.5, V02.51, V02.52, V02.53, V02.54, V02.59, V02.6, V02.60, V02.61, V02.62, V02.69, V02.7, V02.8, V02.9, V03.0, V03.1, V03.2, V03.3, V03.4, V03.5, V03.6, V03.7, V03.8, V03.81, V03.82, V03.89, V03.9, V04.0, V04.1, V04.2, V04.3, V04.4, V04.5, V04.6, V04.7, V04.8, V04.81, V04.82, V04.89, V05.0, V05.1, V05.2, V05.3, V05.4, V05.8, V05.9, V06.0, V06.1, V06.2, V06.3, V06.4, V06.5, V06.6, V06.8, V06.9, V28.6, V71.2, V71.82, V71.83, V73.0, V73.1, V73.2, V73.3, V73.4, V73.5, V73.6, V73.8, V73.81, V73.88, V73.89, V73.9, V73.98, V73.99, V74.0, V74.1, V74.2, V74.3, V74.4, V74.5, V74.6, V74.8, V74.9, V75.0, V75.1, V75.2, V75.3, V75.4, V75.5, V75.6, V75.7, V75.8, V75.9, 780.7, 780.71, 780.79 |
|  | Gastritis and Duodenitis | 535.0, 535.00, 535.01, 535.1, 535.10, 535.11, 535.2, 535.20, 535.21, 535.4, 535.40, 535.41, 535.5, 535.50, 535.51, 535.6, 535.60, 535.61, 535.70, 535.71 | 795.5, 795.51, 795.52, 795.6, 795.79, V01.0, V01.1, V01.2, V01.3, V01.4, V01.5, V01.6, V01.7, V01.71, V01.79, V01.8, V01.81, V01.82, V01.83, V01.84, V01.89, V01.9, V02.0, V02.1, V02.2, V02.3, V02.4, V02.5, V02.51, V02.52, V02.53, V02.54, V02.59, V02.6, V02.60, V02.61, V02.62, V02.69, V02.7, V02.8, V02.9, V03.0, V03.1, V03.2, V03.3, V03.4, V03.5, V03.6, V03.7, V03.8, V03.81, V03.82, V03.89, V03.9, V04.0, V04.1, V04.2, V04.3, V04.4, V04.5, V04.6, V04.7, V04.8, V04.81, V04.82, V04.89, V05.0, V05.1, V05.2, V05.3, V05.4, V05.8, V05.9, V06.0, V06.1, V06.2, V06.3, V06.4, V06.5, V06.6, V06.8, V06.9, V28.6, V71.2, V71.82, V71.83, V73.0, V73.1, V73.2, V73.3, V73.4, V73.5, V73.6, V73.8, V73.81, V73.88, V73.89, V73.9, V73.98, V73.99, V74.0, V74.1, V74.2, V74.3, V74.4, V74.5, V74.6, V74.8, V74.9, V75.0, V75.1, V75.2, V75.3, V75.4, V75.5, V75.6, V75.7, V75.8, V75.9, 535.0, 535.00, 535.01, 535.1, 535.10, 535.11, 535.2, 535.20, 535.21, 535.4, 535.40, 535.41, 535.5, 535.50, 535.51, 535.6, 535.60, 535.61, 535.70, 535.71 |
|  | Essential Hypertension | 401.1, 401.9 | 795.5, 795.51, 795.52, 795.6, 795.79, V01.0, V01.1, V01.2, V01.3, V01.4, V01.5, V01.6, V01.7, V01.71, V01.79, V01.8, V01.81, V01.82, V01.83, V01.84, V01.89, V01.9, V02.0, V02.1, V02.2, V02.3, V02.4, V02.5, V02.51, V02.52, V02.53, V02.54, V02.59, V02.6, V02.60, V02.61, V02.62, V02.69, V02.7, V02.8, V02.9, V03.0, V03.1, V03.2, V03.3, V03.4, V03.5, V03.6, V03.7, V03.8, V03.81, V03.82, V03.89, V03.9, V04.0, V04.1, V04.2, V04.3, V04.4, V04.5, V04.6, V04.7, V04.8, V04.81, V04.82, V04.89, V05.0, V05.1, V05.2, V05.3, V05.4, V05.8, V05.9, V06.0, V06.1, V06.2, V06.3, V06.4, V06.5, V06.6, V06.8, V06.9, V28.6, V71.2, V71.82, V71.83, V73.0, V73.1, V73.2, V73.3, V73.4, V73.5, V73.6, V73.8, V73.81, V73.88, V73.89, V73.9, V73.98, V73.99, V74.0, V74.1, V74.2, V74.3, V74.4, V74.5, V74.6, V74.8, V74.9, V75.0, V75.1, V75.2, V75.3, V75.4, V75.5, V75.6, V75.7, V75.8, V75.9, 401.1, 401.9 |
|  | Esophageal Disorders | 456.1, 456.21, 530.0, 530.1, 530.10, 530.11, 530.12, 530.13, 530.19, 530.2, 530.20, 530.21, 530.3, 530.4, 530.5, 530.6, 530.8, 530.81, 530.83, 530.84, 530.85, 530.89, 530.9 | 795.5, 795.51, 795.52, 795.6, 795.79, V01.0, V01.1, V01.2, V01.3, V01.4, V01.5, V01.6, V01.7, V01.71, V01.79, V01.8, V01.81, V01.82, V01.83, V01.84, V01.89, V01.9, V02.0, V02.1, V02.2, V02.3, V02.4, V02.5, V02.51, V02.52, V02.53, V02.54, V02.59, V02.6, V02.60, V02.61, V02.62, V02.69, V02.7, V02.8, V02.9, V03.0, V03.1, V03.2, V03.3, V03.4, V03.5, V03.6, V03.7, V03.8, V03.81, V03.82, V03.89, V03.9, V04.0, V04.1, V04.2, V04.3, V04.4, V04.5, V04.6, V04.7, V04.8, V04.81, V04.82, V04.89, V05.0, V05.1, V05.2, V05.3, V05.4, V05.8, V05.9, V06.0, V06.1, V06.2, V06.3, V06.4, V06.5, V06.6, V06.8, V06.9, V28.6, V71.2, V71.82, V71.83, V73.0, V73.1, V73.2, V73.3, V73.4, V73.5, V73.6, V73.8, V73.81, V73.88, V73.89, V73.9, V73.98, V73.99, V74.0, V74.1, V74.2, V74.3, V74.4, V74.5, V74.6, V74.8, V74.9, V75.0, V75.1, V75.2, V75.3, V75.4, V75.5, V75.6, V75.7, V75.8, V75.9, 456.1, 456.21, 530.0, 530.1, 530.10, 530.11, 530.12, 530.13, 530.19, 530.2, 530.20, 530.21, 530.3, 530.4, 530.5, 530.6, 530.8, 530.81, 530.83, 530.84, 530.85, 530.89, 530.9 |
|  | Coronary Atherosclerosis and Other Heart Disease | 411.0, 411.1, 411.8, 411.81, 411.89, 412, 413.0, 413.1, 413.9, 414.0, 414.00, 414.01, 414.06, 414.2, 414.3, 414.4, 414.8, 414.9, V45.81, V45.82 | 795.5, 795.51, 795.52, 795.6, 795.79, V01.0, V01.1, V01.2, V01.3, V01.4, V01.5, V01.6, V01.7, V01.71, V01.79, V01.8, V01.81, V01.82, V01.83, V01.84, V01.89, V01.9, V02.0, V02.1, V02.2, V02.3, V02.4, V02.5, V02.51, V02.52, V02.53, V02.54, V02.59, V02.6, V02.60, V02.61, V02.62, V02.69, V02.7, V02.8, V02.9, V03.0, V03.1, V03.2, V03.3, V03.4, V03.5, V03.6, V03.7, V03.8, V03.81, V03.82, V03.89, V03.9, V04.0, V04.1, V04.2, V04.3, V04.4, V04.5, V04.6, V04.7, V04.8, V04.81, V04.82, V04.89, V05.0, V05.1, V05.2, V05.3, V05.4, V05.8, V05.9, V06.0, V06.1, V06.2, V06.3, V06.4, V06.5, V06.6, V06.8, V06.9, V28.6, V71.2, V71.82, V71.83, V73.0, V73.1, V73.2, V73.3, V73.4, V73.5, V73.6, V73.8, V73.81, V73.88, V73.89, V73.9, V73.98, V73.99, V74.0, V74.1, V74.2, V74.3, V74.4, V74.5, V74.6, V74.8, V74.9, V75.0, V75.1, V75.2, V75.3, V75.4, V75.5, V75.6, V75.7, V75.8, V75.9, 411.0, 411.1, 411.8, 411.81, 411.89, 412, 413.0, 413.1, 413.9, 414.0, 414.00, 414.01, 414.06, 414.2, 414.3, 414.4, 414.8, 414.9, V45.81, V45.82 |
|  | Congestive Heart Failure; Non-Hypertensive | 398.91, 428.0, 428.1, 428.20, 428.21, 428.22, 428.23, 428.30, 428.31, 428.32, 428.33, 428.40, 428.41, 428.42, 428.43, 428.9 | 795.5, 795.51, 795.52, 795.6, 795.79, V01.0, V01.1, V01.2, V01.3, V01.4, V01.5, V01.6, V01.7, V01.71, V01.79, V01.8, V01.81, V01.82, V01.83, V01.84, V01.89, V01.9, V02.0, V02.1, V02.2, V02.3, V02.4, V02.5, V02.51, V02.52, V02.53, V02.54, V02.59, V02.6, V02.60, V02.61, V02.62, V02.69, V02.7, V02.8, V02.9, V03.0, V03.1, V03.2, V03.3, V03.4, V03.5, V03.6, V03.7, V03.8, V03.81, V03.82, V03.89, V03.9, V04.0, V04.1, V04.2, V04.3, V04.4, V04.5, V04.6, V04.7, V04.8, V04.81, V04.82, V04.89, V05.0, V05.1, V05.2, V05.3, V05.4, V05.8, V05.9, V06.0, V06.1, V06.2, V06.3, V06.4, V06.5, V06.6, V06.8, V06.9, V28.6, V71.2, V71.82, V71.83, V73.0, V73.1, V73.2, V73.3, V73.4, V73.5, V73.6, V73.8, V73.81, V73.88, V73.89, V73.9, V73.98, V73.99, V74.0, V74.1, V74.2, V74.3, V74.4, V74.5, V74.6, V74.8, V74.9, V75.0, V75.1, V75.2, V75.3, V75.4, V75.5, V75.6, V75.7, V75.8, V75.9, 398.91, 428.0, 428.1, 428.20, 428.21, 428.22, 428.23, 428.30, 428.31, 428.32, 428.33, 428.40, 428.41, 428.42, 428.43, 428.9 |
|  | Conditions Associated with Dizziness or Vertigo | 386.00, 386.01, 386.02, 386.03, 386.04, 386.10, 386.11, 386.12, 386.19, 386.2, 386.30, 386.31, 386.32, 386.33, 386.34, 386.35, 386.40, 386.41, 386.42, 386.43, 386.48, 386.50, 386.51, 386.52, 386.53, 386.54, 386.55, 386.56, 386.58, 386.8, 386.9, 780.4 | 795.5, 795.51, 795.52, 795.6, 795.79, V01.0, V01.1, V01.2, V01.3, V01.4, V01.5, V01.6, V01.7, V01.71, V01.79, V01.8, V01.81, V01.82, V01.83, V01.84, V01.89, V01.9, V02.0, V02.1, V02.2, V02.3, V02.4, V02.5, V02.51, V02.52, V02.53, V02.54, V02.59, V02.6, V02.60, V02.61, V02.62, V02.69, V02.7, V02.8, V02.9, V03.0, V03.1, V03.2, V03.3, V03.4, V03.5, V03.6, V03.7, V03.8, V03.81, V03.82, V03.89, V03.9, V04.0, V04.1, V04.2, V04.3, V04.4, V04.5, V04.6, V04.7, V04.8, V04.81, V04.82, V04.89, V05.0, V05.1, V05.2, V05.3, V05.4, V05.8, V05.9, V06.0, V06.1, V06.2, V06.3, V06.4, V06.5, V06.6, V06.8, V06.9, V28.6, V71.2, V71.82, V71.83, V73.0, V73.1, V73.2, V73.3, V73.4, V73.5, V73.6, V73.8, V73.81, V73.88, V73.89, V73.9, V73.98, V73.99, V74.0, V74.1, V74.2, V74.3, V74.4, V74.5, V74.6, V74.8, V74.9, V75.0, V75.1, V75.2, V75.3, V75.4, V75.5, V75.6, V75.7, V75.8, V75.9, 386.00, 386.01, 386.02, 386.03, 386.04, 386.10, 386.11, 386.12, 386.19, 386.2, 386.30, 386.31, 386.32, 386.33, 386.34, 386.35, 386.40, 386.41, 386.42, 386.43, 386.48, 386.50, 386.51, 386.52, 386.53, 386.54, 386.55, 386.56, 386.58, 386.8, 386.9, 780.4 |
|  | Cardiac Dysrhythmia | 427.0, 427.1, 427.2, 427.31, 427.32, 427.60, 427.61, 427.69, 427.81, 427.89, 427.9, 785.0, 785.1 | 795.5, 795.51, 795.52, 795.6, 795.79, V01.0, V01.1, V01.2, V01.3, V01.4, V01.5, V01.6, V01.7, V01.71, V01.79, V01.8, V01.81, V01.82, V01.83, V01.84, V01.89, V01.9, V02.0, V02.1, V02.2, V02.3, V02.4, V02.5, V02.51, V02.52, V02.53, V02.54, V02.59, V02.6, V02.60, V02.61, V02.62, V02.69, V02.7, V02.8, V02.9, V03.0, V03.1, V03.2, V03.3, V03.4, V03.5, V03.6, V03.7, V03.8, V03.81, V03.82, V03.89, V03.9, V04.0, V04.1, V04.2, V04.3, V04.4, V04.5, V04.6, V04.7, V04.8, V04.81, V04.82, V04.89, V05.0, V05.1, V05.2, V05.3, V05.4, V05.8, V05.9, V06.0, V06.1, V06.2, V06.3, V06.4, V06.5, V06.6, V06.8, V06.9, V28.6, V71.2, V71.82, V71.83, V73.0, V73.1, V73.2, V73.3, V73.4, V73.5, V73.6, V73.8, V73.81, V73.88, V73.89, V73.9, V73.98, V73.99, V74.0, V74.1, V74.2, V74.3, V74.4, V74.5, V74.6, V74.8, V74.9, V75.0, V75.1, V75.2, V75.3, V75.4, V75.5, V75.6, V75.7, V75.8, V75.9, 427.0, 427.1, 427.2, 427.31, 427.32, 427.60, 427.61, 427.69, 427.81, 427.89, 427.9, 785.0, 785.1 |
|  | Abdominal Pain | 789.0, 789.00, 789.01, 789.02, 789.03, 789.04, 789.05, 789.06, 789.07, 789.09, 789.60, 789.61, 789.62, 789.63, 789.64, 789.65, 789.66, 789.67, 789.69 | 795.5, 795.51, 795.52, 795.6, 795.79, V01.0, V01.1, V01.2, V01.3, V01.4, V01.5, V01.6, V01.7, V01.71, V01.79, V01.8, V01.81, V01.82, V01.83, V01.84, V01.89, V01.9, V02.0, V02.1, V02.2, V02.3, V02.4, V02.5, V02.51, V02.52, V02.53, V02.54, V02.59, V02.6, V02.60, V02.61, V02.62, V02.69, V02.7, V02.8, V02.9, V03.0, V03.1, V03.2, V03.3, V03.4, V03.5, V03.6, V03.7, V03.8, V03.81, V03.82, V03.89, V03.9, V04.0, V04.1, V04.2, V04.3, V04.4, V04.5, V04.6, V04.7, V04.8, V04.81, V04.82, V04.89, V05.0, V05.1, V05.2, V05.3, V05.4, V05.8, V05.9, V06.0, V06.1, V06.2, V06.3, V06.4, V06.5, V06.6, V06.8, V06.9, V28.6, V71.2, V71.82, V71.83, V73.0, V73.1, V73.2, V73.3, V73.4, V73.5, V73.6, V73.8, V73.81, V73.88, V73.89, V73.9, V73.98, V73.99, V74.0, V74.1, V74.2, V74.3, V74.4, V74.5, V74.6, V74.8, V74.9, V75.0, V75.1, V75.2, V75.3, V75.4, V75.5, V75.6, V75.7, V75.8, V75.9, 789.0, 789.00, 789.01, 789.02, 789.03, 789.04, 789.05, 789.06, 789.07, 789.09, 789.60, 789.61, 789.62, 789.63, 789.64, 789.65, 789.66, 789.67, 789.69 |
| **Tuberculosis** | Tonsillitis | 463, 474.0, 474.00, 474.01, 474.02, 474.10, 474.11, 475, 474.12, 474.2, 474.8, 474.9 | J03.80, J03.81, J03.90, J03.91, J35.01, J35.02, J35.03, J35.2, J35.3, J35.8, J35.9, J36, J35.1 |
|  | Respiratory Failure | 517.3, 518.81, 518.82, 518.83, 518.84, 799.1 | J96.11, J96.91, R09.01, J80, J96.00, J96.01, J96.02, J96.10, J96.12, J96.20, J96.21, J96.22, J96.90, J96.92 |
|  | Respiratory Cancer | 163.0, 163.1, 163.8, 163.9, 165.0, 165.8, 165.9, 231.1, 231.8, 231.9 | C33, C38.4, C39.0, C39.9, C45.0, D02.1, D02.3, D02.4 |
|  | Pneumonia | 112.4, 114.0, 114.4, 115.05, 115.15, 115.95, 130.4, 136.3, 480.0, 480.1, 480.2, 480.8, 480.9, 481, 482.0, 482.1, 482.2, 482.3, 482.30, 482.31, 482.32, 482.39, 482.4, 482.40, 482.41, 482.42, 482.49, 482.8, 482.81, 482.83, 482.84, 482.89, 482.9, 483, 483.0, 483.1, 483.8, 484.1, 484.3, 484.6, 484.7, 484.8, 485, 486, 513.0, 517.1 | A31.0, A37.01, A37.11, A43.0, A48.1, B25.0, B37.1, B38.0, B38.1, B38.2, B39.0, B39.1, B39.2, B58.3, B59, B77.81, J12.0, J12.1, J12.2, J12.3, J12.89, J12.9, J13, J14, J15.0, J15.1, J15.20, J15.211, J15.212, J15.29, J15.3, J15.4, J15.5, J15.6, J15.7, J15.8, J15.9, J16.0, J16.8, J17, J18.0, J18.1, J18.8, J18.9, J85.1 |
|  | Pleurisy Pneumothorax | 510.0, 511.0, 511.1, 511.8, 511.89, 512.0, 512.8, 512.81, 512.82, 512.83, 512.84, 512.89, 518.1, 518.2, 511.9, 518.0 | J86.0, J93.11, J86.9, J92.0, J92.9, J93.0, J93.12, J93.81, J93.82, J93.83, J94.0, J94.1, J94.2, J94.8, J94.9, J98.19, J98.2, J98.3, R09.1, J90, J91.8, J93.9, J98.11 |
|  | Other Upper-Respiratory Infection | 784.91, 460, 461.0, 461.1, 461.2, 461.3, 461.9, 462, 464.0, 464.00, 464.01, 464.11, 464.20, 464.21, 464.30, 464.31, 464.4, 464.50, 464.51, 465.0, 465.8, 465.9, 473.0, 473.1, 473.2, 473.3, 473.8, 473.9, 464.10 | J00, J01.00, J01.01, J01.10, J01.11, J01.20, J01.21, J01.30, J01.31, J01.40, J01.41, J01.80, J01.81, J01.90, J01.91, J02.0, J02.8, J02.9, J03.00, J03.01, J04.0, J04.10, J04.11, J04.2, J04.30, J04.31, J05.0, J05.10, J05.11, J06.0, J06.9, J32.0, J32.1, J32.2, J32.3, J32.4, J32.8, J32.9, R09.82 |
|  | Other Upper Respiratory Disease | 784.1, 784.41, 784.42, 784.9, 784.99, 472.2, 476.0, 476.1, 478.21, 478.22, 478.24, 478.71, 478.9, 519.2, 472.0, 477.0, 477.2, 477.8, 477.9, 478.1, 478.19, 478.20, 478.29, 478.30, 478.31, 478.32, 478.33, 478.34, 478.4, 478.5, 478.70, 478.74, 478.75, 478.79, 478.8, 519.1, 519.11, 519.19, 519.3, 784.40, 784.49, 784.7, 784.8 | R49.0, R49.8, R49.9, J31.1, J31.2, J39.0, J39.1, J98.09, J98.5, J98.59, R07.0, J30.0, J30.1, J30.2, J30.81, J30.89, J30.9, J31.0, J34.2, J34.89, J37.0, J37.1, J38.00, J38.01, J38.02, J38.1, J38.2, J38.3, J38.4, J38.5, J38.6, J38.7, J39.2, J39.3, J39.8, J39.9, J98.01, J98.51, R04.0, R04.1, R09.81 |
|  | Other Lower Respiratory Disease | 786.02, 786.05, 786.07, 786.2, 786.3, 786.30, 786.4, 786.52, 513.1, 514, 515, 516.0, 516.1, 516.2, 516.3, 516.30, 516.31, 516.32, 516.33, 516.34, 516.35, 516.36, 516.37, 516.4, 516.5, 516.8, 516.9, 517.2, 517.8, 518.3, 518.4, 518.89, 519.4, 519.8, 519.9, 786.00, 786.09, 786.39, 786.9, 793.11, 794.2, 786.6, 786.7, 793.1, 793.19 | R04.2, R05, R06.00, R06.01, R06.02, R06.03, R06.09, R06.2, R06.89, R06.9, R07.1, R07.81, R09.3, J18.2, J22, J85.2, J85.3, J98.9, R06.6, R06.82, J81.0, J81.1, J82, J84.01, J84.02, J84.03, J84.09, J84.10, J84.111, J84.112, J84.113, J84.114, J84.115, J84.116, J84.117, J84.17, J84.2, J84.81, J84.82, J84.89, J84.9, J98.4, J98.6, J98.8, J99, R04.89, R04.9, R09.02, R91.1, R91.8 |
|  | Lung Disease Due to External Agents | 495.0, 495.1, 495.2, 495.3, 495.4, 495.5, 495.6, 495.7, 495.8, 495.9, 500, 501, 502, 503, 504, 505, 506.0, 506.1, 506.2, 506.3, 506.4, 506.9, 507.1, 507.8, 508.0, 508.1, 508.2, 508.8, 508.9 | J60, J61, J62.0, J62.8, J63.0, J63.1, J63.2, J63.3, J63.4, J63.5, J63.6, J64, J66.0, J66.1, J66.2, J66.8, J67.0, J67.1, J67.2, J67.3, J67.4, J67.5, J67.6, J67.7, J67.8, J67.9, J68.0, J68.1, J68.2, J68.3, J68.4, J68.8, J68.9, J69.1, J69.8, J70.0, J70.1, J70.2, J70.3, J70.4, J70.5, J70.8, J70.9 |
|  | Lung Cancer | 162.2, 162.3, 162.4, 162.5, 162.8, 162.9, 209.21 | C34.00, C34.01, C34.02, C34.10, C34.11, C34.12, C34.2, C34.30, C34.31, C34.32, C34.80, C34.81, C34.82, C34.90, C34.91, C34.92, C7A.090, D02.20, D02.21, D02.22 |
|  | Influenza | 487.0, 487.1, 487.8, 488, 488.1, 488.11, 488.12, 488.19, 488.81, 488.82, 488.89 | J09.X1, J09.X2, J09.X3, J09.X9, J10.00, J10.01, J10.08, J10.1, J10.2, J10.89, J11.00, J11.08, J11.1, J11.2, J11.81, J11.82, J11.83, J11.89 |
|  | Hemoptysis | 786.3, 786.30, 786.39 | R04.2, R04.8, R04.89, R04.9 |
|  | Fever | 780.6, 780.60, 780.61 | R50, R50.81, R50.9 |
|  | Cough | 786.2 | R05 |
|  | COPD | 490, 491.0, 491.1, 491.8, 491.9, 491.2, 491.20, 491.21, 491.22, 492.0, 492.8, 494, 494.0, 494.1, 496 | J40, J41.0, J41.1, J42, J44.0, J47.0, J41.8, J43.0, J43.1, J43.2, J43.8, J43.9, J44.1, J44.9, J47.1, J47.9 |
|  | Bronchitis | 466.0, 466.1, 466.11, 466.19 | J20.0, J20.1, J20.2, J20.3, J20.4, J20.5, J20.6, J20.7, J20.8, J20.9, J21.0, J21.1, J21.8, J21.9 |
|  | Asthma | 493.00, 493.01, 493.02, 493.10, 493.11, 493.12, 493.20, 493.21, 493.22, 493.81, 493.82, 493.90, 493.92 | J45.20, J45.21, J45.22, J45.30, J45.31, J45.32, J45.40, J45.41, J45.42, J45.50, J45.51, J45.52, J45.901, J45.902, J45.909, J45.990, J45.991, J45.998 |
|  | Aspiration Pneumonitis | 507 | J69.0 |
|  | Additional Codes | 780.79, 780.8, 783.21, 786.50, 786.51, 786.59, 038.9, 079.99, 310, 340, 391, 599.0, 830, 995.91, 135, 197.0, 212.3, 235.7, 239.1, 289.1, 416.8, 423.9, 428.0, 446.4, 263.9, 276.1, 285.29, 285.9, 288.60, 289.3, 429.3, 782.2, 784.2, 785.0, 785.6, 799.02, 799.4 | R07.2, R07.82, R07.89, R07.9, R53.1, R53.81, R53.83, R61, R63.4, A41.9, B34.9, D14.30, D38.1, D49.1, D86.0, D86.9, I50.9, J85.0, N39.0, D64.9, D72.829, E871, I51.7, R00.0, R22.0, R22.1, R22.2, R59, R59.0, R59.1, R59.9 |

**Supplementary Table 3. Change-point, delay window, and number of missed opportunities by change-point detection approach and disease**

|  | **Stroke** | | | **AMI** | | | **Tuberculosis** | | |
| --- | --- | --- | --- | --- | --- | --- | --- | --- | --- |
|  | ***Linear- Cubic*** | ***CUSUM*** | ***Prediction Bound*** | ***Linear- Cubic*** | ***CUSUM*** | ***Prediction Bound*** | ***Linear- Cubic*** | ***CUSUM*** | ***Prediction Bound*** |
| **Change-point** | 8 | 30 | 39 | 4 | 35 | 40 | 114 | 78 | 74 |
| **Delay Window** | [8, 1] | [30, 1] | [39, 1] | [4, 1] | [35, 1] | [40, 1] | [114, 1] | [78, 1] | [74, 1] |
| **Total Number of Missed Opportunities During Delay Window (% of SSD visits during delay window)** | 28,630 (77.4%) | 40,034 (64.8%) | 41,577 (60.5%) | 57,596 (74.7%) | 102,103 (43.5%) | 103,874 (40.9%) | 6,444 (58.1%) | 6,247 (65.2%) | 6,068 (65.0%) |

**Supplementary Table 4. Assessment of fit by change-point detection approach**

| **Disease** | **Method** | **MSE Before**  **Change-point** | **Mean error X days before change-point** | | | |
| --- | --- | --- | --- | --- | --- | --- |
|  |  |  | ***7-days*** | ***14-Days*** | ***21-Days*** | ***28-Days*** |
| **Stroke** | Linear Cubic | 23319.6 | 533.7 | 310.3 | 197.4 | 121.5 |
|  | CUSUM | 1759.1 | 102.8 | 72.5 | 55.1 | 41.8 |
|  | Prediction Bound | 1311.0 | 60.0 | 42.0 | 28.1 | 16.8 |
| **AMI** | Linear Cubic | 337499.3 | 2193.8 | 1402.4 | 933.1 | 637.6 |
|  | CUSUM | 16108.7 | 255.7 | 177.7 | 132.1 | 105.9 |
|  | Prediction Bound | 15538.4 | 153.6 | 104.3 | 95.5 | 57.2 |
| **Tuberculosis** | Linear Cubic | 44.4 | -2.6 | -0.9 | -0.5 | 0.3 |
|  | CUSUM | 54.1 | 6.8 | 6.7 | 7.5 | 5.3 |
|  | Prediction Bound | 61.1 | 9.9 | 7.9 | 7.5 | 6.2 |

Note: ME = Mean Error; MSE = Mean Squared Error

**Supplementary Table 5. Estimates of the number of missed opportunities used in the simulation models.**

| **Days Prior to Index** | **Stroke** | **AMI** | **Tuberculosis** |
| --- | --- | --- | --- |
| 1 | 12472 | 40871 | 272 |
| 2 | 5350 | 9696 | 247 |
| 3 | 3561 | 6297 | 189 |
| 4 | 2788 | 4902 | 192 |
| 5 | 2322 | 4163 | 155 |
| 6 | 1817 | 3925 | 215 |
| 7 | 1592 | 3747 | 253 |
| 8 | 1320 | 2844 | 191 |
| 9 | 1028 | 2208 | 153 |
| 10 | 837 | 1986 | 137 |
| 11 | 785 | 1775 | 138 |
| 12 | 680 | 1725 | 118 |
| 13 | 692 | 1890 | 164 |
| 14 | 715 | 1896 | 203 |
| 15 | 654 | 1447 | 133 |
| 16 | 415 | 1072 | 106 |
| 17 | 372 | 1083 | 125 |
| 18 | 308 | 925 | 100 |
| 19 | 311 | 912 | 78 |
| 20 | 298 | 1036 | 128 |
| 21 | 359 | 1247 | 162 |
| 22 | 317 | 802 | 142 |
| 23 | 279 | 493 | 94 |
| 24 | 188 | 530 | 77 |
| 25 | 154 | 608 | 65 |
| 26 | 189 | 662 | 100 |
| 27 | 238 | 650 | 90 |
| 28 | 322 | 839 | 123 |
| 29 | 181 | 571 | 102 |
| 30 | 214 | 365 | 80 |
| 31 | 135 | 336 | 75 |
| 32 | 42 | 257 | 72 |
| 33 | 114 | 241 | 71 |
| 34 | 142 | 410 | 80 |
| 35 | 143 | 625 | 96 |
| 36 | 125 | 300 | 85 |
| 37 | 70 | 206 | 63 |
| 38 | 15 | 150 | 52 |
| 39 | 33 | 29 | 43 |
| 40 | - | 153 | 73 |
| 41 | - | - | 78 |
| 42 | - | - | 88 |
| 43 | - | - | 41 |
| 44 | - | - | 49 |
| 45 | - | - | 29 |
| 46 | - | - | 34 |
| 47 | - | - | 25 |
| 48 | - | - | 62 |
| 49 | - | - | 70 |
| 50 | - | - | 57 |
| 51 | - | - | 26 |
| 52 | - | - | 15 |
| 53 | - | - | 38 |
| 54 | - | - | 37 |
| 55 | - | - | 33 |
| 56 | - | - | 63 |
| 57 | - | - | 39 |
| 58 | - | - | 22 |
| 59 | - | - | 31 |
| 60 | - | - | 8 |
| 61 | - | - | 11 |
| 62 | - | - | 26 |
| 63 | - | - | 44 |
| 64 | - | - | 27 |
| 65 | - | - | 16 |
| 66 | - | - | 22 |
| 67 | - | - | 10 |
| 68 | - | - | 15 |
| 69 | - | - | 15 |
| 70 | - | - | 30 |
| 71 | - | - | 30 |
| 72 | - | - | 7 |
| 73 | - | - | 1 |
| 74 | - | - | 10 |
| 75 | - | - | 0 |
| 76 | - | - | 31 |
| 77 | - | - | 27 |
| 78 | - | - | 24 |
| 79 | - | - | 0 |
| 80 | - | - | 2 |
| 81 | - | - | 0 |
| 82 | - | - | 0 |
| 83 | - | - | 9 |
| 84 | - | - | 31 |
| 85 | - | - | 4 |
| 86 | - | - | 0 |
| 87 | - | - | 0 |
| 88 | - | - | 21 |
| 89 | - | - | 1 |
| 90 | - | - | 23 |
| 91 | - | - | 6 |
| 92 | - | - | 3 |
| 93 | - | - | 15 |
| 94 | - | - | 6 |
| 95 | - | - | 0 |
| 96 | - | - | 11 |
| 97 | - | - | 5 |
| 98 | - | - | 24 |
| 99 | - | - | 4 |
| 100 | - | - | 0 |
| 101 | - | - | 3 |
| 102 | - | - | 0 |
| 103 | - | - | 7 |
| 104 | - | - | 0 |
| 105 | - | - | 6 |
| 106 | - | - | 0 |
| 107 | - | - | 0 |
| 108 | - | - | 3 |
| 109 | - | - | 0 |
| 110 | - | - | 0 |
| 111 | - | - | 6 |
| 112 | - | - | 15 |
| 113 | - | - | 6 |
| 114 | - | - | 0 |

**Supplementary Table 6. Expanded Simulation results for stroke**

|  | **Algorithm 1** | **Algorithm 2 (alpha = 0)** | **Algorithm 2 (alpha = 1)** |
| --- | --- | --- | --- |
| **Number of Missed Opportunities (n 95% CI))** |  |  |  |
| Outpatient | 26042 (62.6%) [CI: 25951 -26144 (62.4 - 62.9%)] | 26211 (63.0%) [CI: 26109 - 26317 (62.8 - 63.3%)] | 25579 (61.5%) [CI: 25502 - 25656 (61.3 - 61.7%)] |
| Inpatient | 1815 (4.4%) [CI: 1765 - 1861 (4.2 - 4.5%)] | 1771 (4.3%) [CI: 1725 - 1818 (4.1 - 4.4%)] | 1871 (4.5%) [CI: 1831 - 1915 (4.4 - 4.6%)] |
| ED | 13720 (33.0%) [CI: 13631 -13807 (32.8 - 33.2%)] | 13595 (32.7%) [CI: 13500 - 13688 (32.5 - 32.9%)] | 14127 (34%) [CI: 14054 - 14203 (33.8 - 34.2%)] |
| Total | 41577 | 41577 | 41577 |
| **Number of Patients Experiencing Missed Opportunities (n (% of all patients) [95% CI])** |  |  |  |
| 0 | 340347 (92.5%) [340258 - 340437 (92.5 - 92.6%)] | 342268 (93.1%) [342169 - 342368 (93.0 - 93.1%)] | 337272 (91.7%) [337215 - 337326 (91.7 - 91.7%)] |
| ≥ 1 | 27421 (7.5%) [27331 - 27510 (7.4 - 7.5%)] | 25500 (6.9%) [25400 - 25599 (6.9 - 7.0%)] | 30496 (8.3%) [30442 - 30553 (8.3 - 8.3%)] |
| ≥ 2 | 8748 (2.4%) [8679 - 8819 (2.4 - 2.4%)] | 8680 (2.4%) [8606 - 8751 (2.3 - 2.4%)] | 8148 (2.2%) [8091 - 8204 (2.2 - 2.2%)] |
| ≥ 3 | 3122 (0.8%) [3074 - 3175 (0.8 - 0.9%)] | 3526 (1.0%) [3474 - 3580 (0.9 - 1.0%)] | 2138 (0.6%) [2103 - 2174 (0.6 - 0.6%)] |
| ≥ 4 | 1239 (0.3%) [1201 - 1277 (0.3 - 0.3%)] | 1653 (0.4%) [1613 - 1696 (0.4 - 0.5%)] | 572 (0.2%) [545 - 599 (0.1 - 0.2%)] |
| ≥ 5 | 539 (0.1%) [512 - 567 (0.1 - 0.2%)] | 861 (0.2%) [830 - 893 (0.2 - 0.2%)] | 163 (0.0%) [147 - 180 (0.0 - 0.0%)] |
| Mean - Overall | 1.52 [1.51 - 1.52] | 1.63 [1.62 - 1.64] | 1.36 [1.36 - 1.37] |
| Median - Overall | 1 [1 - 1] | 1 [1 - 1] | 1 [1 - 1] |
| Mean - Outpatient | 0.95 [0.95 - 0.95] | 1.03 [1.02 - 1.03] | 0.84 [0.84 - 0.84] |
| Median - Outpatient | 1 [1 - 1] | 1 [1 - 1] | 1 [1 - 1] |
| Mean - Inpatient | 0.07 [0.06 - 0.07] | 0.07 [0.07 - 0.07] | 0.06 [0.06 - 0.06] |
| Median - Inpatient | 0 [0 - 0] | 0 [0 - 0] | 0 [0 - 0] |
| Mean - ED | 0.50 [0.50 - 0.50] | 0.53 [0.53 - 0.54] | 0.46 [0.46 - 0.47] |
| Median - ED | 0 [0 - 0] | 0 [0 - 0] | 0 [0 - 0] |
| **Durations of Delays (n (% of patients experiencing missed opportunities) [95% CI])** |  |  |  |
| ≥ 1 days | 27421 (100.0%) [27331 - 27510 (100.0 - 100.0%)] | 25500 (100.0%) [25400 - 25599 (100.0 - 100.0%)] | 30496 (100.0%) [30442 - 30553 (100.0 - 100.0%)] |
| ≥ 3 days | 16047 (58.5%) [15966 - 16126 (58.3 - 58.7%)] | 14107 (55.3%) [14006 - 14196 (55.1 - 55.5%)] | 18841 (61.8%) [18792 - 18891 (61.7 - 61.9%)] |
| ≥ 7 days | 9899 (36.1%) [9827 - 9967 (35.9 - 36.3%)] | 8189 (32.1%) [8106 - 8266 (31.8 - 32.4%)] | 11879 (39.0%) [11835 - 11920 (38.8 - 39.1%)] |
| ≥ 11 days | 6775 (24.7%) [6717 - 6833 (24.5 - 24.9%)] | 5405 (21.2%) [5328 - 5474 (20.9 - 21.4%)] | 7882 (25.8%) [7846 - 7919 (25.8 - 25.9%)] |
| ≥ 15 days | 4714 (17.2%) [4667 - 4760 (17.0 - 17.4%)] | 3680 (14.4%) [3615 - 3739 (14.2 - 14.6%)] | 5316 (17.4%) [5287 - 5345 (17.3 - 17.5%)] |
| ≥ 19 days | 3366 (12.3%) [3330 - 3402 (12.1 - 12.4%)] | 2614 (10.2%) [2559 - 2666 (10.0 - 10.4%)] | 3709 (12.2%) [3687 - 3730 (12.1 - 12.2%)] |
| ≥ 22 days | 2580 (9.4%) [2549 - 2609 (9.3 - 9.5%)] | 1834 (7.2%) [1789 - 1877 (7.0 - 7.4%)] | 2493 (8.2%) [2476 - 2510 (8.1 - 8.2%)] |
| ≥ 26 days | 1793 (6.5%) [1770 - 1816 (6.5 - 6.6%)] | 1361 (5.3%) [1326 - 1395 (5.2 - 5.5%)] | 1717 (5.6%) [1704 - 1730 (5.6 - 5.7%)] |
| Mean - Among Delayed | 7.41 [7.38 - 7.44] | 6.72 [6.67 - 6.76] | 7.64 [7.63 - 7.66] |
| Median - Among Delayed | 4 [4 - 4] | 3 [3 - 3] | 4 [4 - 4] |
| Mean - Everyone Included | 0.55 [0.55 - 0.56] | 0.47 [0.46 - 0.47] | 0.63 [0.63 - 0.64] |
| Median - Everyone Included | 0 [0 - 0] | 0 [0 - 0] | 0 [0 - 0] |

**Supplementary Table 7. Expanded simulation results for AMI**

|  | **Algorithm 1** | **Algorithm 2 (alpha = 0)** | **Algorithm 2 (alpha = 1)** |
| --- | --- | --- | --- |
| **Number of Missed Opportunities (n 95% CI))** |  |  |  |
| Outpatient | 78772 (75.8%) [CI: 78610 - 78930 (75.7 – 76.0%)] | 79367 (76.4%) [CI: 79200 - 79529 (76.2 - 76.6%)] | 77457 (74.6%) [CI: 77321 - 77595 (74.4 - 74.7%)] |
| Inpatient | 6115 (5.9%) [CI: 6019 - 6214 (5.8 - 6.0%)] | 6089 (5.9%) [CI: 5991 - 6185 (5.8 - 6.0%)] | 6131 (5.9%) [CI: 6037 - 6216 (5.8 - 6.0%)] |
| ED | 18987 (18.3%) [CI: 18853 - 19128 (18.1 - 18.4%)] | 18418 (17.7%) [CI: 18280 - 18567 (17.6 - 17.9%)] | 20286 (19.5%) [CI: 20169 - 20397 (19.4 - 19.6%)] |
| Total | 103874 | 103874 | 103874 |
| **Number of Patients Experiencing Missed Opportunities (n (% of all patients) [95% CI])** |  |  |  |
| 0 | 293111 (81.5%) [292943 - 293284 (81.5 - 81.6%)] | 302032 (84.0%) [301825 - 302233 (83.9 - 84.0%)] | 282986 (78.7%) [282877 - 283093 (78.7 - 78.7%)] |
| ≥ 1 | 66514 (18.5%) [66341 - 66682 (18.4 - 18.5%)] | 57593 (16.0%) [57392 - 57800 (16.0 - 16.1%)] | 76639 (21.3%) [76532 - 76748 (21.3 - 21.3%)] |
| ≥ 2 | 24689 (6.9%) [24563 - 24828 (6.8 - 6.9%)] | 23912 (6.6%) [23781 - 24053 (6.6 - 6.7%)] | 21332 (5.9%) [21233 - 21425 (5.9 - 6.0%)] |
| ≥ 3 | 8158 (2.3%) [8051 - 8257 (2.2 - 2.3%)] | 9951 (2.8%) [9847 - 10054 (2.7 - 2.8%)] | 4796 (1.3%) [4736 - 4859 (1.3 - 1.4%)] |
| ≥ 4 | 2748 (0.8%) [2680 - 2813 (0.7 - 0.8%)] | 4784 (1.3%) [4706 - 4861 (1.3 - 1.4%)] | 896 (0.2%) [863 - 931 (0.2 - 0.3%)] |
| ≥ 5 | 1011 (0.3%) [964 - 1054 (0.3 - 0.3%)] | 2616 (0.7%) [2555 - 2681 (0.7 - 0.7%)] | 171 (0.0%) [154 - 189 (0.0 - 0.1%)] |
| Mean - Overall | 1.56 [1.56 - 1.57] | 1.80 [1.80 - 1.81] | 1.36 [1.35 - 1.36] |
| Median - Overall | 1 [1 - 1] | 1 [1 - 1] | 1 [1 - 1] |
| Mean - Outpatient | 1.18 [1.18 - 1.19] | 1.38 [1.37 - 1.38] | 1.01 [1.01 - 1.01] |
| Median - Outpatient | 1 [1 - 1] | 1 [1 - 1] | 1 [1 - 1] |
| Mean - Inpatient | 0.09 [0.09 - 0.09] | 0.11 [0.10 - 0.11] | 0.08 [0.08 - 0.08] |
| Median - Inpatient | 0 [0 - 0] | 0 [0 - 0] | 0 [0 - 0] |
| Mean - ED | 0.29 [0.28 - 0.29] | 0.32 [0.32 - 0.32] | 0.26 [0.26 - 0.27] |
| Median - ED | 0 [0 - 0] | 0 [0 - 0] | 0 [0 - 0] |
| **Durations of Delays (n (% of patients experiencing missed opportunities) [95% CI])** |  |  |  |
| ≥ 1 days | 66514 (100.0%) [66341 - 66682 (100.0 - 100.0%)] | 57593 (100.0%) [57392 - 57800 (100.0 - 100.0%)] | 76639 (100.0%) [76532 - 76748 (100.0 - 100.0%)] |
| ≥ 3 days | 37711 (56.7%) [37568 - 37868 (56.5 - 56.8%)] | 24563 (42.6%) [24381 - 24744 (42.4 - 42.9%)] | 42329 (55.2%) [42252 - 42407 (55.1 - 55.3%)] |
| ≥ 7 days | 26262 (39.5%) [26134 - 26387 (39.3 - 39.6%)] | 16031 (27.8%) [15877 - 16200 (27.6 - 28.1%)] | 28472 (37.2%) [28406 - 28537 (37.1 - 37.2%)] |
| ≥ 11 days | 19042 (28.6%) [18940 - 19146 (28.5 - 28.8%)] | 11673 (20.3%) [11532 - 11810 (20.0 - 20.5%)] | 20523 (26.8%) [20471 - 20576 (26.7 - 26.8%)] |
| ≥ 15 days | 13710 (20.6%) [13635 - 13792 (20.5 - 20.7%)] | 8072 (14.0%) [7949 - 8191 (13.8 - 14.2%)] | 14053 (18.3%) [14013 - 14089 (18.3 - 18.4%)] |
| ≥ 19 days | 10131 (15.2%) [10071 - 10192 (15.1 - 15.3%)] | 6224 (10.8%) [6114 - 6331 (10.6 - 11.0%)] | 10228 (13.3%) [10196 - 10257 (13.3 - 13.4%)] |
| ≥ 23 days | 6812 (10.2%) [6768 - 6857 (10.2 - 10.3%)] | 4316 (7.5%) [4227 - 4408 (7.3 - 7.6%)] | 6790 (8.9%) [6766 - 6812 (8.8 - 8.9%)] |
| ≥ 27 days | 4803 (7.2%) [4771 - 4835 (7.2 - 7.3%)] | 3091 (5.4%) [3024 - 3163 (5.3 - 5.5%)] | 4403 (5.7%) [4384 - 4420 (5.7 - 5.8%)] |
| Mean - Among Delayed | 8.10 [8.08 - 8.12] | 6.69 [6.64 - 6.73] | 8.21 [8.20 - 8.22] |
| Median - Among Delayed | 4 [4 - 4] | 2 [2 - 2] | 5 [5 - 5] |
| Mean - Everyone Included | 1.50 [1.49 - 1.50] | 1.07 [1.06 - 1.08] | 1.75 [1.75 - 1.75] |
| Median - Everyone Included | 0 [0 - 0] | 0 [0 - 0] | 0 [0 - 0] |

**Supplementary Table 8. Expanded simulation results for tuberculosis**

|  | **Algorithm 1** | **Algorithm 2 (alpha = 0)** | **Algorithm 2 (alpha = 1)** |
| --- | --- | --- | --- |
| **Number of Missed Opportunities (n 95% CI))** |  |  |  |
| Outpatient | 5750 (89.2%) [CI: 5724 - 5778 (88.8 - 89.7%)] | 5816 (90.3%) [CI: 5788 - 5846 (89.8 - 90.7%)] | 5708 (88.6%) [CI: 5684 - 5733 (88.2 - 89%)] |
| Inpatient | 324 (5.0%) [CI: 304 - 342 (4.7 - 5.3%)] | 315 (4.9%) [CI: 297 - 333 (4.6 - 5.2%)] | 327 (5.1%) [CI: 310 - 343 (4.8 - 5.3%)] |
| ED | 370 (5.7%) [CI: 348 - 390 (5.4 - 6.1%)] | 313 (4.9%) [CI: 291 - 334 (4.5 - 5.2%)] | 409 (6.3%) [CI: 391 - 425 (6.1 - 6.6%)] |
| Total | 6444 | 6444 | 6444 |
| **Number of Patients Experiencing Missed Opportunities (n (% of all patients) [95% CI])** |  |  |  |
| 0 | 454 (21.9%) [438 - 470 (21.1 - 22.7%)] | 748 (36.1%) [725 - 771 (35.0 - 37.2%)] | 367 (17.7%) [361 - 374 (17.4 - 18.0%)] |
| ≥ 1 | 1619 (78.1%) [1603 - 1635 (77.3 - 78.9%)] | 1325 (63.9%) [1302 - 1348 (62.8 - 65.0%)] | 1706 (82.3%) [1699 - 1712 (82.0 - 82.6%)] |
| ≥ 2 | 1309 (63.2%) [1290 - 1329 (62.2 - 64.1%)] | 1071 (51.7%) [1049 - 1092 (50.6 - 52.7%)] | 1391 (67.1%) [1375 - 1408 (66.3 - 67.9%)] |
| ≥ 3 | 1011 (48.8%) [992 - 1032 (47.9 - 49.8%)] | 861 (41.5%) [841 - 880 (40.6 - 42.5%)] | 1062 (51.2%) [1044 - 1081 (50.4 - 52.1%)] |
| ≥ 4 | 756 (36.5%) [736 - 775 (35.5 - 37.4%)] | 695 (33.5%) [675 - 714 (32.6 - 34.4%)] | 765 (36.9%) [747 - 784 (36.0 - 37.8%)] |
| ≥ 5 | 548 (26.4%) [529 - 566 (25.5 - 27.3%)] | 555 (26.8%) [537 - 574 (25.9 - 27.7%)] | 530 (25.6%) [513 - 547 (24.7 - 26.4%)] |
| Mean - Overall | 3.98 [3.94 - 4.02] | 4.87 [4.78 - 4.95] | 3.78 [3.76 - 3.79] |
| Median - Overall | 3 [3 - 3] | 4 [4 - 4] | 3 [3 - 3] |
| Mean - Outpatient | 3.55 [3.51 - 3.59] | 4.39 [4.31 - 4.47] | 3.35 [3.33 - 3.37] |
| Median - Outpatient | 3 [3 - 3] | 4 [3 - 3] | 3 [3 - 3] |
| Mean - Inpatient | 0.20 [0.19 - 0.21] | 0.24 [0.22 - 0.25] | 0.19 [0.18 - 0.20] |
| Median - Inpatient | 0 [0 - 0] | 0 [0 - 0] | 0 [0 - 0] |
| Mean - ED | 0.23 [0.22 - 0.24] | 0.24 [0.22 - 0.25] | 0.24 [0.23 - 0.25] |
| Median - ED | 0 [0 - 0] | 0 [0 - 0] | 0 [0 - 0] |
| **Durations of Delays (n (% of patients experiencing missed opportunities) [95% CI])** |  |  |  |
| ≥ 1 days | 1619 (100.0%) [1603 - 1635 (100.0 - 100.0%)] | 1325 (100.0%) [1302 - 1348 (100.0 - 100.0%)] | 1706 (100.0%) [1699 - 1712 (100.0 - 100.0%)] |
| ≥ 11 days | 1355 (83.7%) [1336 - 1374 (82.7 - 84.6%)] | 1024 (77.3%) [1001 - 1048 (75.6 - 79.0%)] | 1489 (87.3%) [1481 - 1498 (86.9 - 87.7%)] |
| ≥ 22 days | 1099 (67.9%) [1080 - 1120 (66.8 - 69.0%)] | 776 (58.6%) [754 - 801 (56.8 - 60.5%)] | 1267 (74.3%) [1257 - 1277 (73.8 - 74.8%)] |
| ≥ 33 days | 853 (52.7%) [833 - 871 (51.5 - 53.8%)] | 552 (41.7%) [529 - 575 (39.9 - 43.5%)] | 1026 (60.1%) [1015 - 1036 (59.6 - 60.7%)] |
| ≥ 45 days | 623 (38.5%) [603 - 643 (37.3 - 39.7%)] | 387 (29.2%) [368 - 407 (27.8 - 30.9%)] | 820 (48.1%) [809 - 831 (47.5 - 48.7%)] |
| ≥ 56 days | 459 (28.4%) [441 - 476 (27.3 - 29.3%)] | 261 (19.7%) [245 - 278 (18.4 - 21.0%)] | 579 (33.9%) [569 - 589 (33.4 - 34.5%)] |
| ≥ 67 days | 298 (18.4%) [285 - 312 (17.6 - 19.2%)] | 186 (14.1%) [172 - 201 (12.9 - 15.2%)] | 371 (21.7%) [363 - 379 (21.3 - 22.2%)] |
| ≥ 79 days | 173 (10.7%) [163 - 182 (10.1 - 11.2%)] | 122 (9.2%) [111 - 134 (8.4 - 10.2%)] | 197 (11.6%) [191 - 202 (11.2 - 11.9%)] |
| ≥ 90 days | 124 (7.7%) [117 - 131 (7.2 - 8.1%)] | 75 (5.7%) [67 - 84 (5.0 - 6.3%)] | 116 (6.8%) [112 - 119 (6.6 - 7.0%)] |
| Mean - Among Delayed | 39.86 [39.31 - 40.37] | 34.35 [33.52 - 35.15] | 44.47 [44.16 - 44.75] |
| Median - Among Delayed | 36 [34 - 36] | 28 [26 - 28] | 43 [42 - 43] |
| Mean - Everyone Included | 31.13 [30.64 - 31.59] | 21.95 [21.45 - 22.45] | 36.59 [36.30 - 36.85] |
| Median - Everyone Included | 25 [23 - 25] | 10 [9 - 11] | 33 [32 - 33] |

**Supplementary Table 9. Sensitivity analysis using all visits - simulation parameters and results of change-point analysis.**

|  | **Stroke** | **AMI** | **Tuberculosis** |
| --- | --- | --- | --- |
| **Change-point Model** | Prediction Bound | Prediction Bound | Linear Cubic |
| **Change-point** | 31 | 31 | 125 |
| **Delay Window** | [31, 1] | [31, 1] | [125, 1] |
| **Total Number of Missed Opportunities During Delay Window (% of all visits during delay window)** | 214,316 (22.7%) | 170,058 (22.9%) | 9,851 (39.4%) |

**Supplementary Table 10. Selected Simulation Results for sensitivity analysis using**

|  | **Stroke** | **AMI** | **Tuberculosis** |
| --- | --- | --- | --- |
| **Change Point (Start of diagnostic opportunity Window** | 31 | 31 | 125 |
| **Total Number of Missed Opportunities During Delay Window (% of SSD visits during delay window)** | 214,316 (22.75%) | 170,058 (22.94%) | 9,851 (39.41%) |
| **Percent of Missed opportunities in outpatient settings** |  |  |  |
| Algorithm 1 | 85.51 (85.40 - 85.62) | 85.86 (85.75 - 85.97) | 92.73 (92.37 - 93.08) |
| Algorithm 2 | 86.79 (86.66 - 86.91) | 86.79 (86.65 - 86.92) | 93.39 (93.02 - 93.78) |
| Algorithm 3 | 83.71 (83.61 - 83.80) | 84.43 (84.32 - 84.52) | 92.26 (91.95 - 92.60) |
| **Percent of missed opportunities in inpatient settings** |  |  |  |
| Algorithm 1 | 2.55 (2.50 - 2.61) | 2.61 (2.55 - 2.67) | 2.85 (2.62 - 3.09) |
| Algorithm 2 | 2.48 (2.43 - 2.53) | 2.57 (2.52 - 2.63) | 2.82 (2.62 - 3.01) |
| Algorithm 3 | 2.54 (2.49 - 2.59) | 2.65 (2.60 - 2.71) | 2.84 (2.64 - 3.06) |
| **Percent of Missed opportunities in ED settings** |  |  |  |
| Algorithm 1 | 11.94 (11.84 - 12.04) | 11.52 (11.43 - 11.63) | 4.42 (4.16 - 4.70) |
| Algorithm 2 | 10.73 (10.62 - 10.85) | 10.64 (10.52 - 10.76) | 3.79 (3.48 - 4.09) |
| Algorithm 3 | 13.75 (13.66 - 13.84) | 12.92 (12.84 - 13.01) | 4.89 (4.65 - 5.15) |
| **Percent of Patients Experiencing Missed Opportunities (95% CI)** |  |  |  |
| Algorithm 1 | 33.92 (33.84 - 33.99) | 28.88 (28.82 - 28.95) | 90.49 (89.77 - 91.17) |
| Algorithm 2 | 22.43 (22.34 - 22.51) | 20.36 (20.29 - 20.44) | 62.45 (61.07 - 63.97) |
| Algorithm 3 | 44.24 (44.20 - 44.29) | 36.48 (36.43 - 36.52) | 94.56 (94.36 - 94.79) |
| **Mean Number of Missed opportunities among patients missed (95% CI)** |  |  |  |
| Algorithm 1 | 1.72 (1.71 - 1.72) | 1.64 (1.63 - 1.64) | 5.25 (5.21 - 5.29) |
| Algorithm 2 | 2.60 (2.59 - 2.61) | 2.32 (2.31 - 2.33) | 7.61 (7.43 - 7.78) |
| Algorithm 3 | 1.32 (1.32 - 1.32) | 1.30 (1.29 - 1.30) | 5.03 (5.01 - 5.04) |
| **Mean duration (days) of missed opportunities among patients missed (95% CI)** |  |  |  |
| Algorithm 1 | 9.47 (9.45 - 9.49) | 8.44 (8.42 - 8.46) | 52.31 (51.60 - 53.00) |
| Algorithm 2 | 6.79 (6.75 - 6.83) | 5.85 (5.81 - 5.89) | 38.65 (37.35 - 39.90) |
| Algorithm 3 | 9.18 (9.17 - 9.18) | 8.22 (8.22 - 8.23) | 61.53 (61.18 - 61.88) |

**Supplementary Figure 1. Change-point detection approach results for stroke**

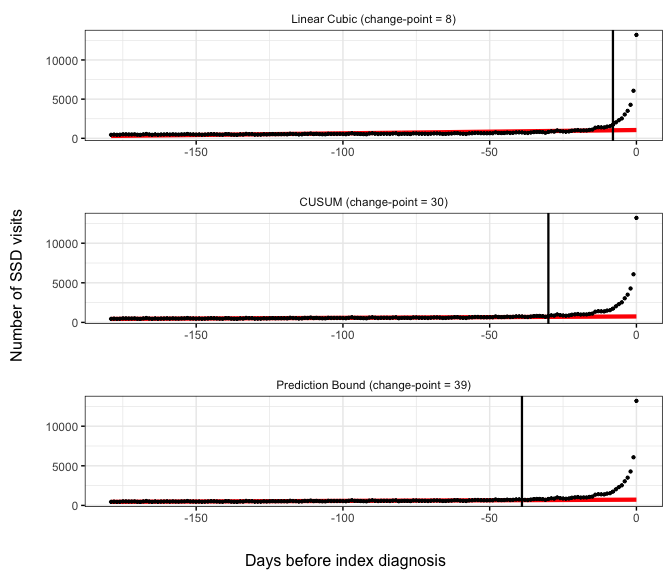

**Supplementary Figure 2: Change-point detection approach results for AMI**

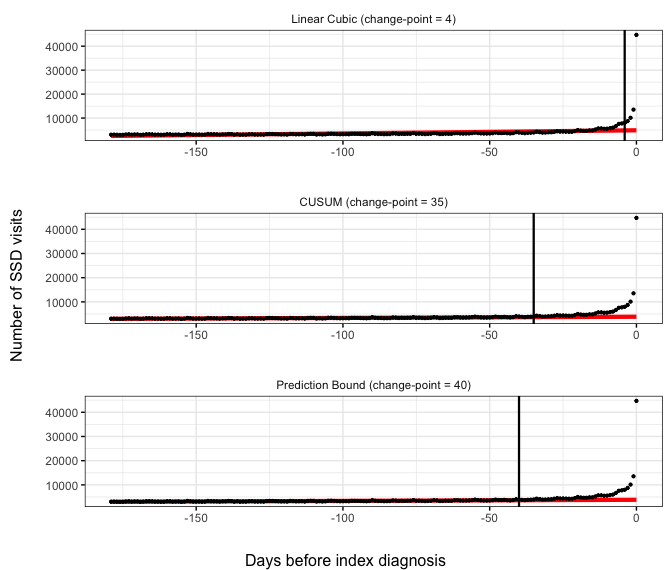

**Supplementary Figure 3. Change-point detection approach results for tuberculosis**

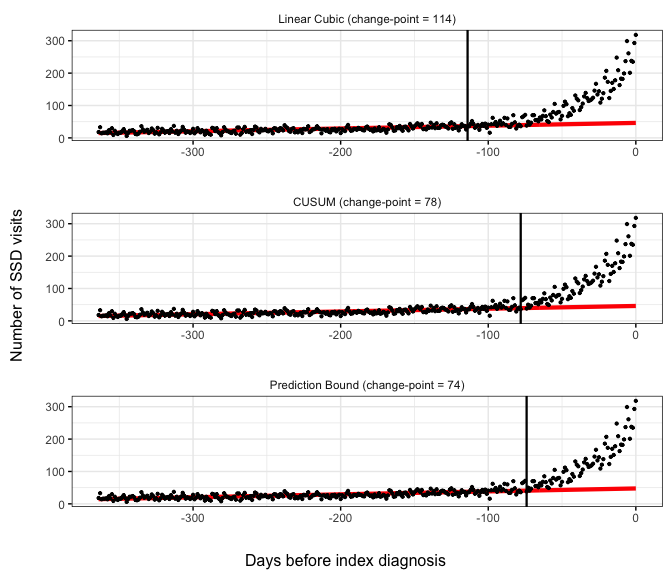
